# Dynamical Systems Modeling of Early-Term Immune Reconstitution with Different Anti-Thymocyte Globulin (ATG) Administration Schedules in Allogeneic Stem Cell Transplantation

**DOI:** 10.1101/2021.05.21.21257336

**Authors:** Viktoriya Zelikson, Gary Simmons, Natasha Raman, Elizabeth Krieger, Anatevka Rebiero, Kelly Hawks, May Aziz, Catherine Roberts, Jason Reed, Ronald Gress, Amir Toor

## Abstract

Alloreactivity forms the basis of allogeneic hematopoietic cell transplantation (HCT), with donor derived T cell response to recipient antigens mediating clinical responses either in part or entirely. These encompass the different manifestations of graft vs. host disease (GVHD), infection risk as well as disease response. Whilst the latter is contingent upon disease biology and thus may be less predictable, the former two are more likely to be directly proportional to the magnitude of donor derived T cell recovery. Herein we explore the quantitative aspects of immune cell recovery following allogeneic HCT and clinical outcomes in two cohorts of HLA matched allograft recipients who received rabbit anti-thymocyte globulin (ATG) on different schedules (days -9 to -7 vs. -3 to -1). Monocyte as well as donor derived T cell (ddCD3) recovery was superior in those given ATG early in their course (days -9/-7). This difference was related to a more rapid rate of ddCD3 recovery, largely driven by CD3+/8+ cells in the first month following transplantation. Early monocyte recovery was associated with later T cell recovery, improved survival, and less chronic GVHD. In contrast rapid and early ddCD3 expansion out of proportion to monocyte recovery was associated with a high likelihood of acute GVHD and poor survival. This analytic methodology demonstrates that modelling ‘early-term immune reconstitution’ following HCT yields insights that may be useful in management of post-transplant immunosuppression and adaptive cellular therapy to optimize clinical outcomes.

**Highlights:** ATG given early (day -9 to -7) prior to HCT leads to improved survival and non-relapse mortality. Improved monocyte counts as well as donor-derived T cell counts are seen in recipients of early ATG. Improved clinical outcomes are observed in patients with enhanced monocyte and T cell recovery after HCT.

## Introduction

Adequate immune reconstitution following allogeneic hematopoietic cell transplantation (HCT) is critical to optimize clinical outcomes, particularly relapse, infection and graft versus host disease (GVHD). Recovery of adaptive immunity is crucial, as it represents the critical mechanism of protection against a variety of viral and fungal pathogens, as well as ensure relapse protection in the long run. Therefore, early T cell reconstitution is crucial to the long-term success of HCT. This is because of the versatility provided by the T cell receptor mediated identification of a wide variety of pathogen and tumor associated epitopes. However, among other factors, T cell recovery also carries with it the risk of GVHD through minor histocompatibility antigen (mHA) recognition.

A number of studies evaluating lymphocyte counts^1^, total T cell and T cell subset recovery at different time points post-transplant have established this paradigm^2^. However, numeric T cell recovery occurs as a continuous function of time, and the risk of transplant related adverse outcomes such as viral infections, GVHD, and relapse varies over the duration of the post-transplant course. In part this is due to variability of T cell recovery over this period. It is therefore important to understand the relationship between clinical events and the magnitude of T cell recovery over time in individual transplant recipients^3^. Further complicating matters is the critical dependence of T cells on antigen presenting cells (represented in the early post-transplant circulation by monocytes), which trigger their antigen specific responses^4^. Developing an understanding of the relationship between the recovery of these populations as a function of time in individual HCT recipients may therefore enhance the understanding of why post-transplant outcomes vary across patient populations, since in individual transplant donor-recipient pairs, HLA and mHA background is quite varied^5, 6^.

In this paper, the relationship between monocyte, T cell and T cell subset recovery over time and their interaction, described using the phrase, early-term immune reconstitution (ETIR), is examined using simple mathematical methods that characterize these interactions as *dynamical systems*. Such systems may be described using differential equations, such as, donor derived T cells proliferating as a logistic function of time with an early exponential growth phase, followed by a later plateau^7^. T cell growth is preceded by an earlier proliferation of monocytes. The magnitude of growth in these populations is variable in different patients, therefore in this paper these parameters were analyzed in individuals. All patients examined received T cell replete grafts and were given rabbit anti-thymocyte globulin (ATG) for in vivo T cell depletion for GVHD prophylaxis. ATG has long been used for in vivo T cell depletion of allografts and is associated with a reduction in the risk for chronic GVHD developing in a number of studies^8, 9^. This risk reduction is associated with increased risk of opportunistic infections, particularly Epstein-Barr Virus associated lymphoproliferative disorder^10^. The effect of ATG on transplant outcomes is dependent on both dose given and schedule of administration^11, 12, 13, 14^. The differential effects observed with ATG dose and schedule variation will logically be dependent on the rate of T cell reconstitution that follows such regimens. In this paper outcomes following two different schedules of ATG administration are reported. Patients were treated with the same pretransplant conditioning regimen and supportive care. ETIR was studied as a function of time in the two cohorts of patients and its impact on clinical outcomes determined.

## Methods

### Patient characteristics and conditioning regimen

The records of 64 consecutive patients transplanted between 2012-2020, using a conditioning regimen of fludarabine and melphalan, were reviewed in a retrospective study approved by the Institutional Review Board at Virginia Commonwealth University Health System. Donors and recipients were matched at 8/8 or 7/8, HLA-A, B, C and DRB1 loci; for unrelated donors, the HLA matching was at the allele level. Conditioning regimen consisted of fludarabine 30 mg/m^2^ administered intravenously (IV) daily from day -6 to -3 and melphalan 140 mg/m^2^ IV on day -2. Total ATG (Thymoglobulin, Sanofi, Bridgewater, NJ) doses utilized were 5 mg/kg for HLA-matched unrelated donor (MUD) and 3 mg/kg for HLA-matched related donor (MRD) recipients. Study patients were given ATG in divided doses from day -9 to -7 (ATG -9/-7 cohort) and their outcomes were compared with a historical cohort given ATG from day -3 to -1 (ATG -3/-1 cohort). Tacrolimus was commenced on either day -3 or -2 and given for approximately 6 months post-transplant for GVHD prophylaxis in MUD recipients, and cyclosporine administered similarly was utilized for MRD transplants. Methotrexate (5mg/m^2^ on day 1, 3, 6 and 11) or mycophenolate mofetil (15 mg/kg every 12 hours from day 0 to 30) were administered along with calcineurin inhibitors. Filgrastim (GCSF 5 μg/kg/day) was given for post-transplant myeloid recovery. Herpes virus infection prophylaxis was administered, with acyclovir 400 mg twice daily, along with cytomegalovirus and Epstein-Barr virus monitoring.

### Early-Term Immune Reconstitution

Circulating monocyte, CD3+, CD3+/8+, CD3+/4+, CD3-/56+ and CD19+ cell counts were determined on days 30, 60, and 90 post-transplant as a measure of early-term immune reconstitution (ETIR). Donor T cell chimerism was measured at these time points utilizing PCR for short tandem repeats on cell populations isolated using anti-CD3 immunomagnetic beads. T cell chimerism, expressed as a fraction was multiplied with the T cell and T cell subset counts (measured within one week of each other), to determine donor-derived T cell (ddCD3) counts. T cell reconstitution kinetics were studied by plotting ddCD3 cell counts for each patient as a function of time, starting with a count of 0 on day 0 (date of transplant) and plotting the counts through day 90. This was followed by fitting a cubic polynomial curve (*f(x)=ax*^*3*^*+bx*^*2*^*+cx+d*) to these data using MS Excel (Microsoft Corporation, Redmond, WA). In this equation, *x* represents the time *t* post-transplant, and *f(x)* the T cell counts at time *t*. The derivative of the cubic equation (*dy/dx=3ax*^*2*^*+2bx+c*) represents an estimate of the rate of change of ddCD3 growth (*dT/dt*) at specific time points. Examples are given depicting this analysis for two patients (**Supplementary Figure 1**). The derivatives for ddCD3 (*dT/dt*) curves were calculated for days 15, 45, and 80 following HCT.

### T cell and monocyte interaction analysis

To understand the interaction between recovering monocytes and T cells, these cell populations at different time points were regarded as vector components in a Euclidean *immune phase-space*, together giving the vector 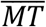. Variation in magnitude of the two cell components over time, change the overall vector magnitude and angle providing a composite estimate of the interaction between these two cell populations and a measure of immune recovery. To accomplish this, monocyte counts on a given day were graphed on the *x*-axis, against ddCD3 cell count on the same day on the *y*-axis, with magnitude and angle of the resulting vector as its properties (**Supplementary Figure 2**).

### Statistical Analysis

Clinical outcomes studied were, overall survival (OS), relapse free survival (RFS), relapse, non-relapse mortality (NRM), aGVHD grades 2-4 and grades 3-4, and cGVHD moderate to severe and severe. Relapsing patients were censored in the NRM analysis at the time of relapse and *vice versa*. Acute and chronic GVHD were determined and graded based on NIH consensus criteria. Those who experienced relapse or received donor lymphocyte infusion (DLI) prior to GVHD diagnosis were censored in the GVHD analysis. Clinical outcomes were studied using Cox proportional hazards modeling using both unadjusted and adjusted analyses. Variables included in the multivariate analysis for clinical outcomes included were, age of recipient (continuous variable), recipient sex, type of disease (myeloid vs lymphoid), at risk status for CMV (defined as IgM or IgG seropositive donor or recipient), donor type (MUD vs MRD), HLA match (either 8/8 or lower), Karnofsky performance status at transplant (either ≥ 90 or below). Cumulative incidence curves were used to compare relapse, NRM, and chronic GVHD.

For immune cell analysis, Wilcoxon-Rank Sum test was used when comparing cell count recovery by cohort. For determining the prognostic impact of ETIR, the two ATG cohorts were combined and cell counts analyzed as categorical variables using cut offs based on ROC-AUC analysis for each cell type, for each time point measured, with OS as the outcome of interest. Univariate and multivariate Cox proportional hazards analysis were performed to assess impact on clinical outcomes. Multivariate analysis for immune cell parameters included type of disease, at risk status for CMV, donor type, HLA match, and ATG timing (ATG -9/-7 vs. ATG -3/-1). This cutoff was applied to calculate Cox proportional hazards for OS, relapse, NRM and GVHD. Statistical analyses were performed using the R Studio (Interface)-V 1.3.1093,2009-202 RStudio, PBC & R-platform V 4.0.3 (2020-10-10).

## Results

### Clinical outcomes with ATG administration early or late in conditioning

Patient characteristics are given in Table 1. The groups were largely comprised of unrelated donor transplant recipients and were comparable, except for a larger proportion of myeloid malignancies in the ATG -9/-7 group. Median follow up in the ATG -9/-7 cohort was 15 months, while it was 47 months in the historical ATG -3/-1 cohort. In unadjusted analysis there was a trend toward improved overall survival in the ATG -9/-7 cohort (Figure 1A). When adjusted for donor type, HLA match grade, disease type, patient age and sex, CMV risk, and KPS, the survival difference between ATG -9/-7 and ATG -3/-1 arms was significant (HR=0.25, p=0.027), and likely attributable to reduced NRM in the ATG -9/-7 cohort (HR=0.14, p=0.027) (Figure 1D & Supplementary Table 1). No difference was observed in relapse (HR=0.77, p=0.73) or RFS (HR=0.40, p=0.11) (Figure 1B & C), or in acute or chronic GVHD (Supplementary Table 1).

**Table 1.** Demographic characteristics of the patients in the two cohorts.

| N | ATG -9/-7 | ATG -3/-1 | P-value |
| --- | --- | --- | --- |
|  | 36 | 28 |  |
| Age (Median) | 56 (27-69) | 57 (17-67) | 0.20 |
| Karnofsky PS (90-100) | 17 (57%) | 18 (67%) | 0.59 |
| Transplant Type (MRD) | 7 (19%) | 10 (36%) | 0.17 |
| GVHD Ppx: MTX+Tacrolimus | 22 | 12 | 0.20 |
| MMF+Cyclosporin | 9 | 10 | 0.41 |
| MTX+Cyclosporin | 4 | 4 | 0.72 |
| MMF+Tacrolimus | 1 | 1 | 1.00 |
| HLA Match (8/8, 10/10) | 28 (78%) | 22 (79%) | 1.00 |
| Disease Type (Myeloid) | 30 (83%)<sup>1</sup> | 10 (36%)<sup>2</sup> | **0.0002** |
| Patient Sex (F) | 19 (53%) | 7 (25%) | **0.04** |
| CD34+ cell dose (mean) (E6/kg) | 4.81 ± 1.16 | 4.87 ± 1.43 | 0.86 |

Diagnoses: 1- ALL (2), AML (13), CML (1), MDS (8), MF (8), MM (1), NHL (1), T cell ALL (2); 2- ALL (1), AML (4), CLL (2), CMML (1), CTCL (1), MF (5), MM (1), NL (10), PCL (1), T cell ALL (2).

**Figure 1A & B:**
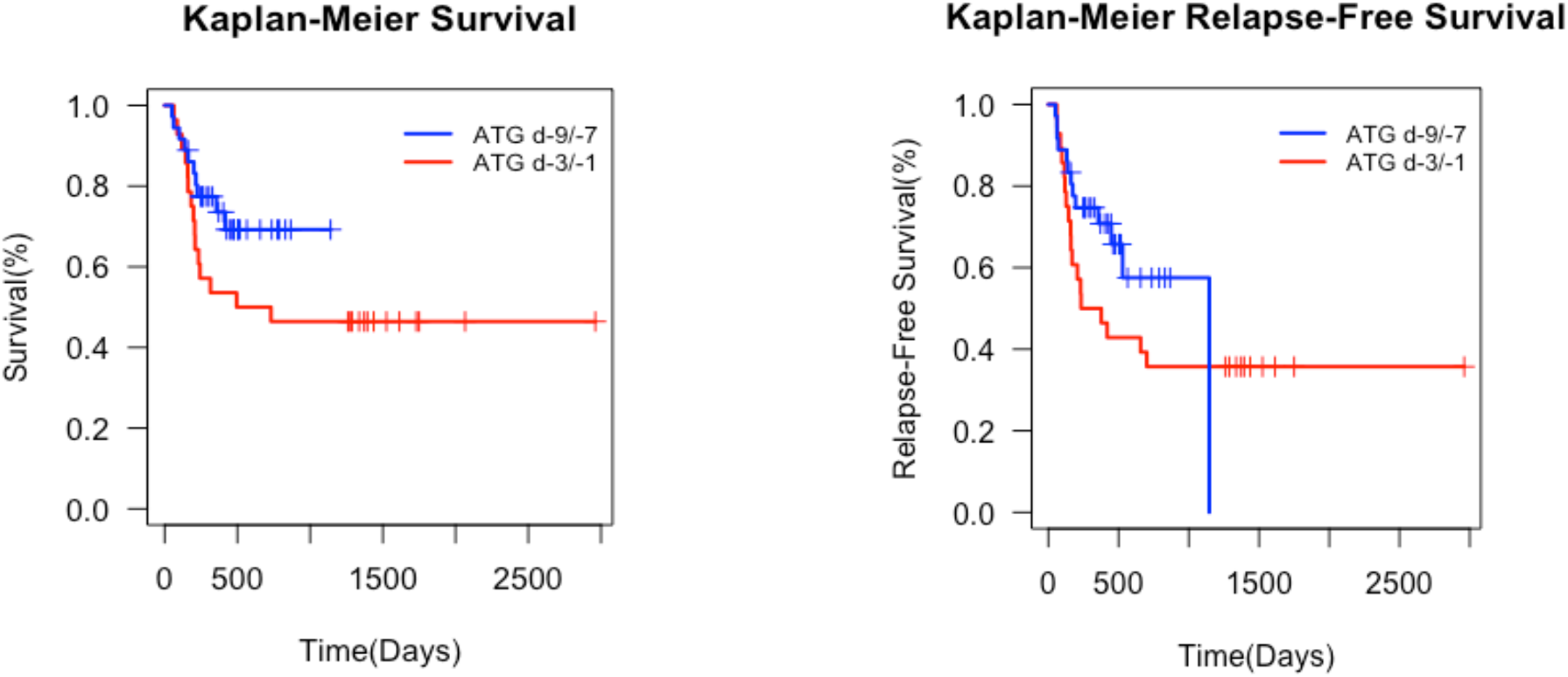
Unadjusted survival (Log Rank p=0.05) and relapse free survival (P=0.2) following HCT in the study, ATG -9/-7 and historical control, ATG -3/-1 cohorts, unadjusted.

**Figure 1C & D.**
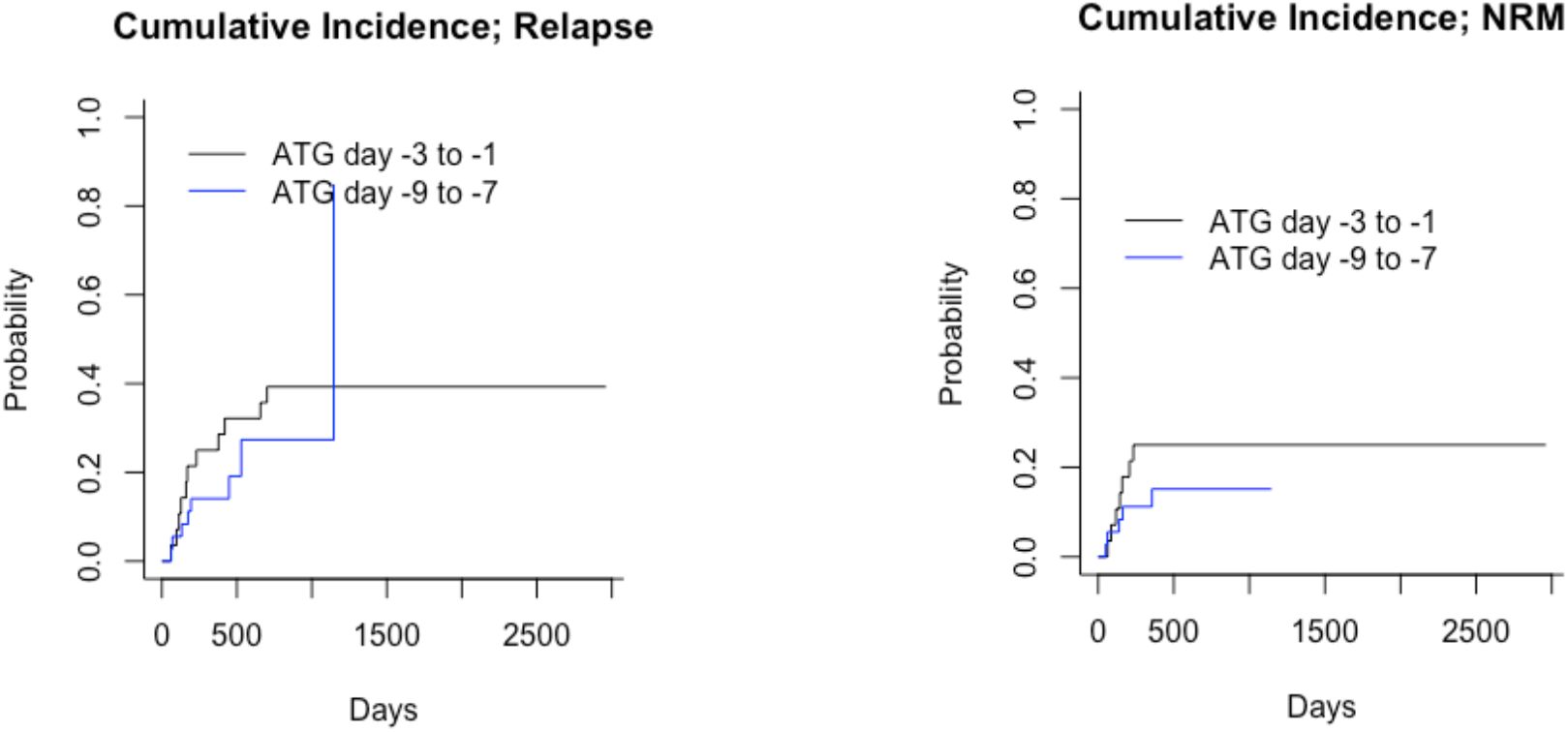
Unadjusted cumulative incidence for relapse (Gray’s test P=0.53) and NRM (P=0.3), unadjusted, for relapse and 0.3 for NRM.

Therapy was tolerated well, with two instances of day 100 treatment-related mortality in each group, due to sepsis (2) in the ATG -9/-7 group, and sepsis (1) and liver failure (1) in the ATG -3/-1 group. There was no difference in acute GVHD (grade 2-4, HR=1.75, p=0.35; grade 3-4, HR=0.27, p=0.23) and chronic GVHD (moderate-severe, HR=1.47, p=0.69; severe, HR=0.54, p=0.62) (Supplementary Figure 3). DLI were utilized for mixed chimerism, relapse, or persistent disease; ATG day -9/-7 (8%) and ATG day - 3/-1(14%) (P=NS). Epstein Barr Virus and CMV reactivation were also similar between the groups (Supplementary Table 2). Of note, viral titers became undetectable in the ATG -9/-7 group in almost half the time compared to the ATG -3/-1 cohort, consistent with superior functional immune recovery in this group.

### Early-term Immune Reconstitution and ATG schedule

To determine the impact of ATG timing on ETIR, immune cell subset recovery and donor-recipient chimerism were compared between the two groups. There was no difference in donor chimerism between the groups (Supplementary Table 3). The ATG -9/-7 group consistently showed superior monocyte recovery throughout the first 90 days following transplantation (Supplementary Table 4). The ddCD3 cell subsets also showed statistically significant divergence by day 60, with ATG -9/-7 group having greater magnitude (Figure 2). The NK cell and CD19+ cell populations showed no difference in recovery between the groups for the time points studied. Early monocyte recovery had a significant impact on later T cell growth (Supplementary figure 5), with monocyte counts on day 30 correlating with ddCD3 cell count on day 60 (Srho: 0.4293, p=0.001), when data from the two cohorts was combined.

**Figure 2.**
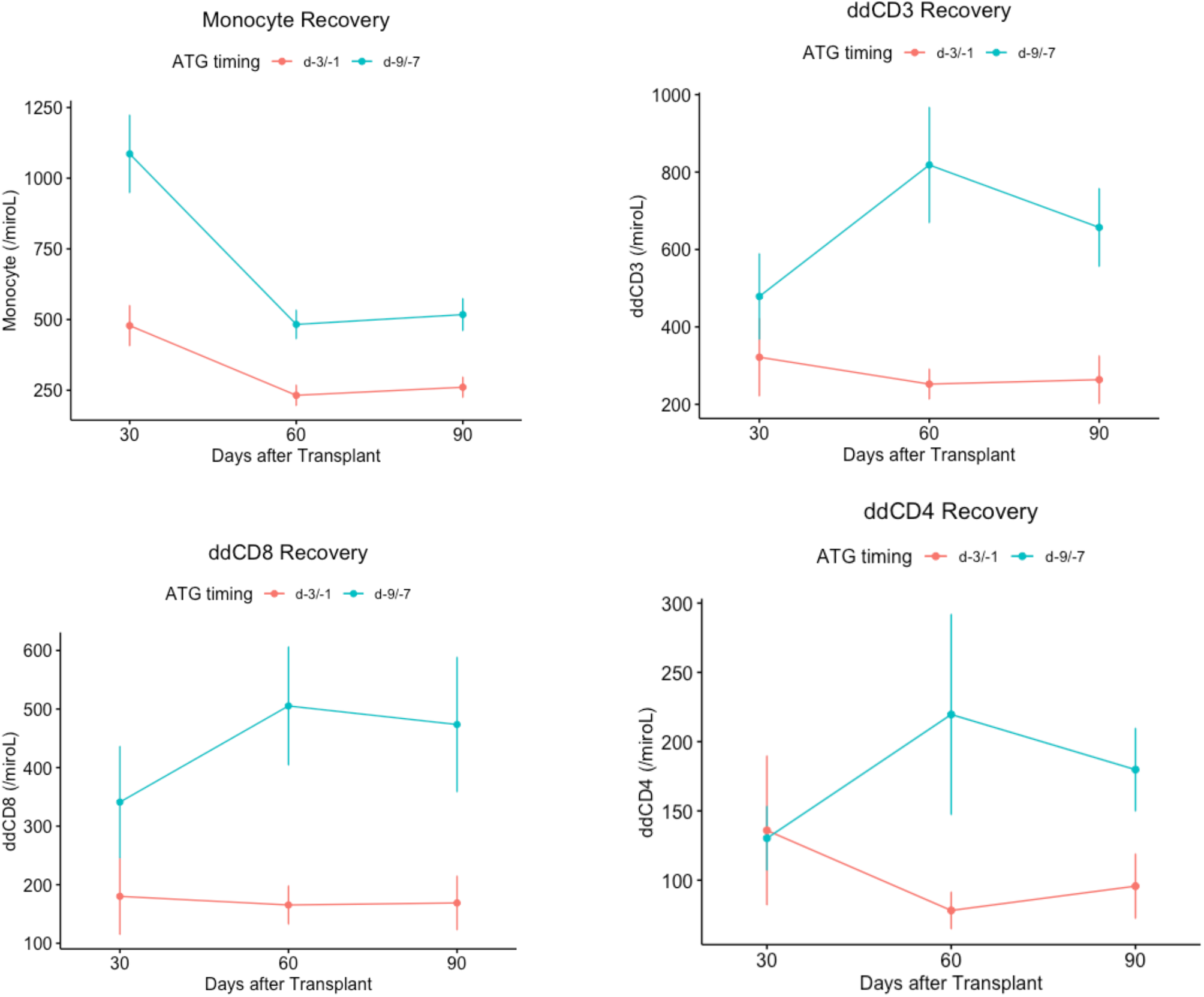
Average immune cell reconstitution on day 30, 60, and 90 post HCT comparing the ATG -9/-7 to ATG -3/-1 cohorts (Mean ± S.E.).

### Early-term Immune Reconstitution and Clinical Outcomes

Monocytes and ddCD3 cell count recovery were analyzed for their impact on clinical outcome. To determine the effect of ETIR on clinical outcomes, data from the two cohorts were combined. The optimal cut-offs for analyzing the impact of monocytes on outcomes were determined utilizing ROC-AUC analysis. The monocyte counts associated with OS on days 30, 60 and 90 were 650/mm^3^, 250/mm^3^, and 50/mm^3^ respectively (Supplementary Figure 5). Univariate and multivariate analysis showed that monocyte count >650/mm^3^ on day 30 was associated with improved OS (univariate HR 0.37, p=0.016; multivariate HR 0.27, p=0.018), as was monocyte count of >250/mm^3^ on day 60 (univariate HR 0.27, p=0.004; multivariate HR 0.34, p=0.056). High monocyte counts were also associated with lower chronic GVHD risk and NRM risk (Figure 3 & Supplementary Table 5A). Similar ROC-AUC analyses for ddCD3+ cell count yielded optimal values of 580/mm^3^, 204/mm^3^, and 278/mm^3^ respectively by day 30, 60 and 90 following HCT. Notably, ddCD3 count >580/mm^3^ on day 30 was associated with increased risk of aGVHD grade 2-4 (univariate HR 3.75, p=0.008; multivariate HR 5.06, p=0.005), and aGVHD grade 3-4 (univariate HR 5.59, p=0.02; multivariate HR 6.3132, p=0.03). A ddCD3 count >204/mm^3^ on day 60 was associated with reduced risk for relapse (univariate HR 0.23, p=0.009; multivariate HR 0.17, p=0.005) (Figure 3 & Supplementary Table 5B).

**Figure 3.**
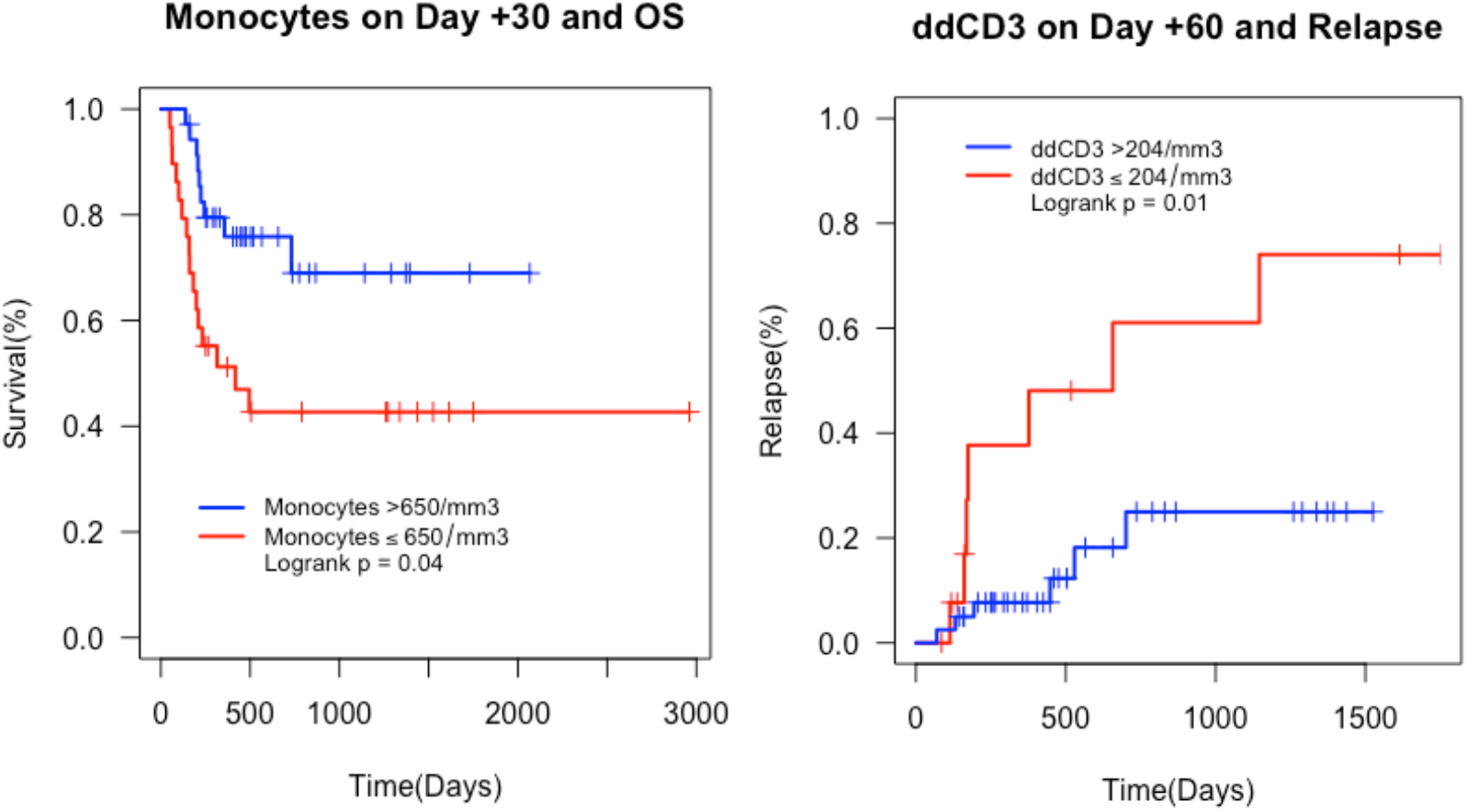
KM curves displaying association of immune cells and outcome.

### T cell Growth Kinetics, ATG schedule and clinical outcomes

The rate of change of ddCD3 was calculated to study the effect of T cell growth kinetics on clinical outcomes. Discrete values of the derivative were calculated for day 15, 45, and 80. There was a significant difference in the *calculated* rate of change (*dT/dt*) on day 15 between the ATG -9/-7 and ATG -3/-1 cohort (16.71 cells.mL^−1^.day^−1^ vs. 8.16, P=0.03, Wilcoxon Rank) (Supplementary Table 6) (Figure 4). This suggests that the different ATG schedule impacts the rate of T cell recovery very early following transplantation, probably driven by monocyte secreted cytokines, and this may contribute to the aforementioned superior T cell recovery observed at the succeeding time points.

**Figure 4.**
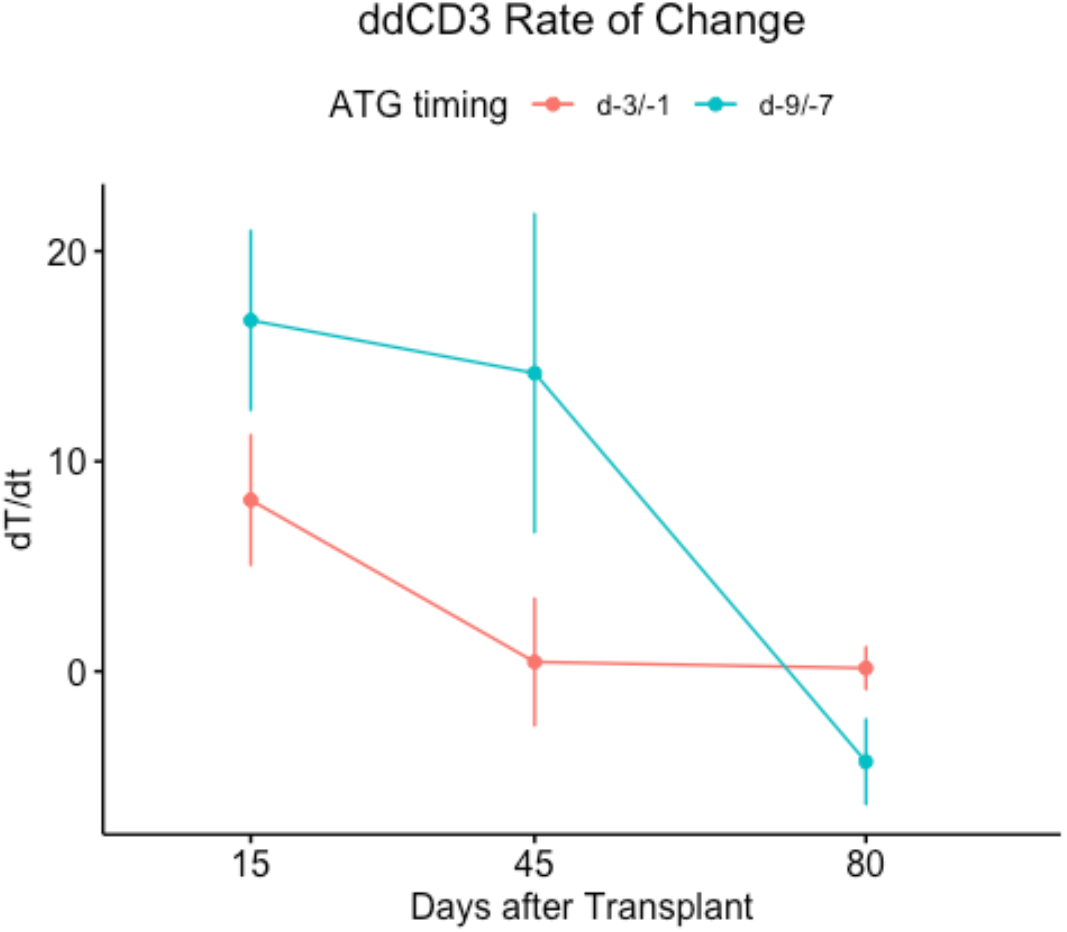
Comparing average ddCD3 rate of change on d15, 45, and 80 between the two cohorts (Mean ± S.E.).

Clinical outcome correlations for ddCD3 kinetics (*dT/dt*) were determined, once again utilizing ROC-AUC analysis for OS, which were optimal at 18.43 cells.mL.^−1^.day^−1^, 0.17, and 4.81 for the time points studied (Supplementary Figure 5). Consistent with the adverse impact of high donor derived T cell counts at day 30 post HCT, there was a significant effect of *dT/dt* >18.43 cells.mL^−1^.day^−1^ on day 15 on both, increased risk of mortality (univariate HR 8.94, p=0.02; bivariate corrected for ATG schedule HR 14.72, p=0.01), as well as aGVHD grade 2-4 (HR 3.23, p=0.048; multivariate HR 4.98, p=0.04). Conversely, consistent with the benefit associated with ddCD3 counts at day 60 post-transplant, there was a significant effect of *dT/dt* >0 cells.mL^−1^.day^−1^ on day 45, with reduced risk of mortality observed (univariate HR 0.09, p=0.03; bivariate corrected for ATG schedule HR 0.1, p=0.04) (Supplementary Table 7), suggesting that ongoing expansion of donor derived T cells beyond day 30 following HCT may confer clinical benefit.

### Monocyte-ddCD3 cell vectors, ATG schedule and clinical outcomes

To examine the interaction of innate (antigen presenting cells) and adaptive (T cells) immune systems, immune reconstitution vectors were constructed using monocyte and ddCD3 cell recovery data (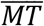). The vector magnitude was greater at all the times examined for the ATG -9/-7 cohort, consistent with superior immune cell recovery. The angle of this vector however was not different at any time between the two cohorts, suggesting the relative magnitude of recovery of the two cell populations was similar in both populations (Figure 5 & Supplementary Table 8).

**Figure 5.**
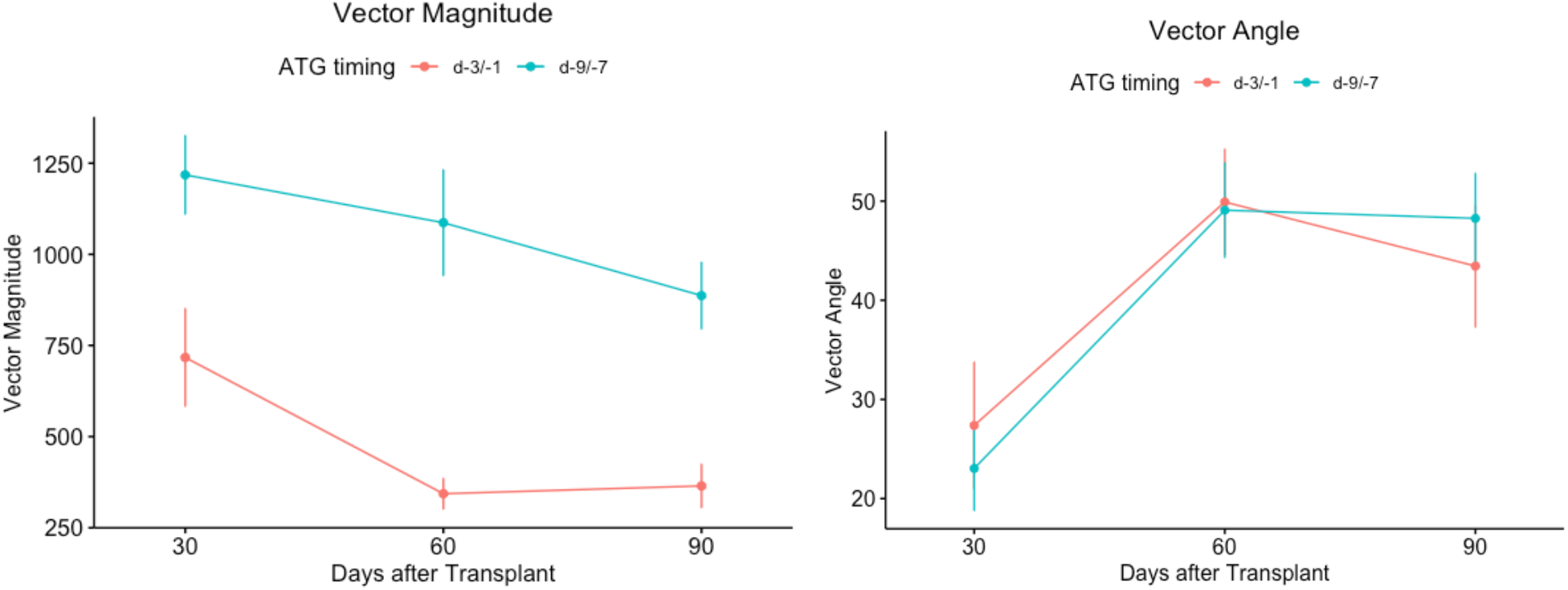
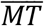 vector magnitude and angle over time in the two patient cohorts (Mean ± S.E.).

Cutoffs for optimal clinical outcome associations for the 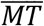 vectors were established through ROC-AUC analysis using OS, and were 955 cells/mm^3^, 619, and 643 for vector magnitude on days 30, 60, and 90 respectively (Supplementary Figure 5). Consistent with the survival benefit observed with T cell and monocyte recovery at day 60, there was a significant difference found with higher vector magnitudes at this time (>619) and improved OS (univariate HR 0.09, p=0.02; multivariate HR 0.08, p=0.03) (Supplementary Table 9). Cut offs for the angle of the vector were established in the same manner, and were 35°, 50°, and 55° for day 30, 60, and 90 respectively. There was a significant difference with an angle >34.65° on day 30 and increased risk of overall mortality (univariate HR 8.15, p=0.001; multivariate HR 10, p=0.007), aGVHD grade 2-4 (univariate HR 3.96, p=0.006; multivariate HR 4.5, p=0.006) and grade 3-4 (univariate HR 11.37, p=0.004; multivariate HR 16.38, p=0.01), as well as cGVHD, both moderate-severe (univariate HR 7.24, p=0.005; multivariate HR 9.15, p=0.006) and severe (univariate HR 175.96, p=0.004) (Supplementary Table 9). These were all associated with greater NRM in these individuals (univariate HR 10, p=0.008; multivariate HR 12, p=0.019). These findings are consistent with the effects of higher *dT/dt* at the early day 15 time point, and high ddCD3, as well as relatively low monocyte counts at this time, pointing to possible rapid, perhaps oligoclonal expansion of alloreactive oligoclonal T cell populations in patients who eventually develop severe GVHD with its adverse impact on survival.

## Discussion

In this paper early-term immune reconstitution (ETIR) following HCT is studied in patients with hematological malignancies, transplanted using two different schedules of ATG. Clinical outcomes are used as a metric for restoration of functional immunity. It is observed that ETIR was more robust, and faster in the patients administered ATG early on (day -9 through -7) in the conditioning regimen. In this small, admittedly varied cohort of patients examined, the related metrics of monocyte and T cell recovery were associated with superior clinical outcomes. This study supports the notion that immune recovery post-transplant functions as a dynamical system, which may be mathematically modeled. This means that a measured intervention (ATG schedule change), produces a measurable impact on a critical variable (ETIR), which may in turn influence the clinical outcomes in a predictable manner (survival, GVHD).

In this study, recipients of ATG early in the course of conditioning had superior monocyte count and a higher rate of donor derived T cell recovery in the first weeks following transplant, which translated to higher T cell counts in the subsequent months. There are several observations to be made which represent areas of future investigative interest. First, robust monocyte recovery was uniformly associated with an improved outlook, with respect to survival and chronic GVHD. Second, higher than average donor derived T cell counts in the first 30 days and very rapid rate of recovery (*dT/dt*) were associated with acute GVHD and diminished survival prospects. However, in the ensuing weeks, likely, as the initial cytokine mediated proliferative surge ebbed, donor derived T cells being above a modest threshold were protective from relapse and associated with improved survival. Finally, when the relative proportion of monocytes and T cells, favored the latter very early on (high 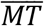 vector angle at day 30), increased alloreactivity with acute and chronic GVHD and worse survival was seen, as noted above, however the salutary effect of vector magnitude mitigated this effect by the second month. These observations suggest that a very early burst of expansion in donor derived T cells accompanied by poor monocyte recovery, is associated with GVHD and impaired survival, while a more tempered growth in accompaniment of robust innate immune recovery (represented by monocytes) leads to improved outcomes (Figure 6). The former scenario may represent a situation where effector memory T cells in the graft encounter disparate antigens and proliferate, whilst the latter may represent polyclonal T cell reconstitution. This creates a picture of a dynamic process unfolding over time as T cells interact with antigen presenting cells, leading to very different outcomes during different time windows in the early-term following transplantation. Further, this process was influenced in a predictable manner by the ATG schedule, with a longer time between ATG and hematopoietic cell infusion resulting in more robust cellular immune recovery.

**Figure 6.**
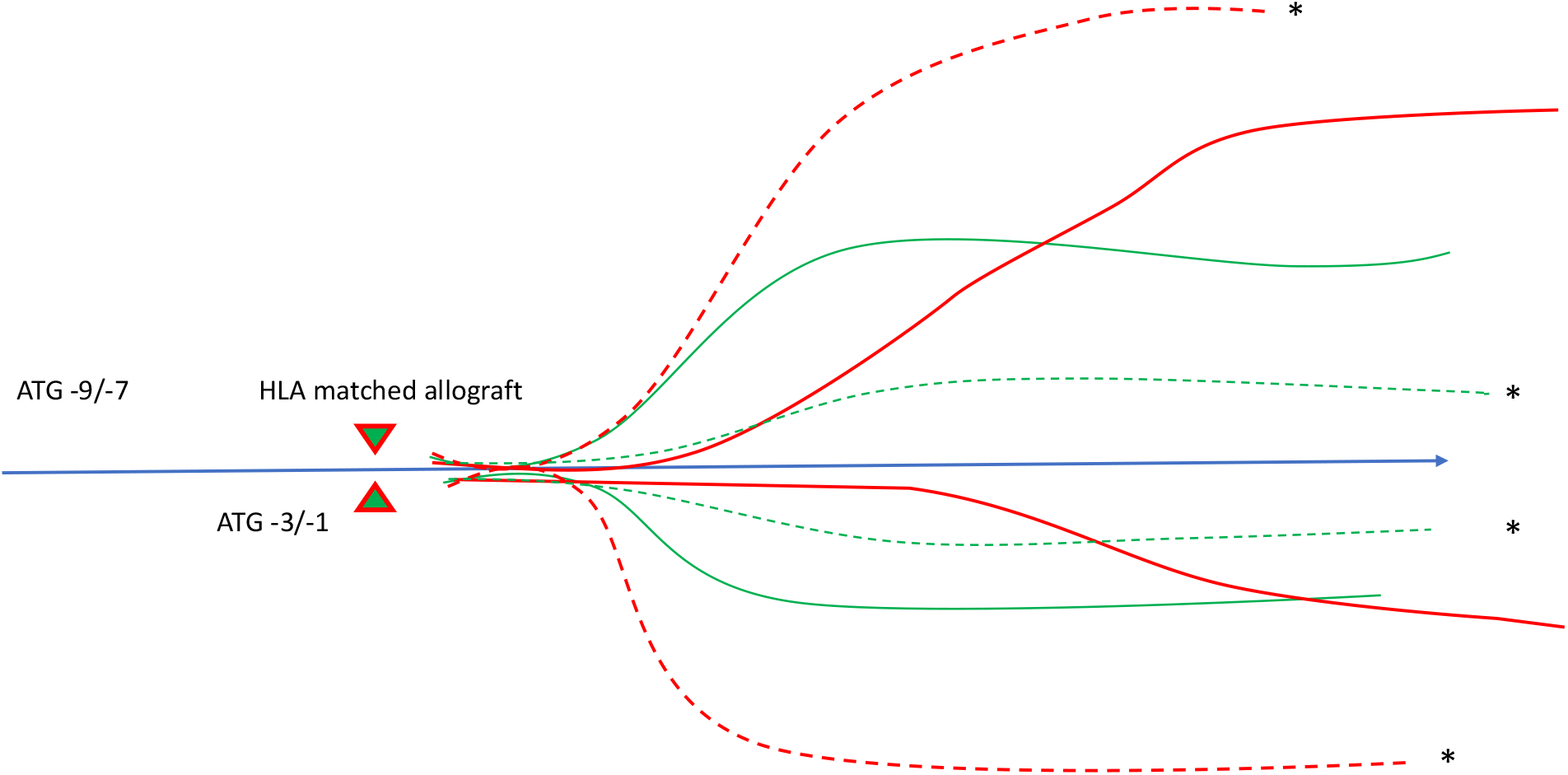
Model of how ATG schedule impacts monocyte (green lines) and T cell (red lines) recovery. Dashed lines indicate rapid uncontrolled T cell growth and poor monocyte recovery, leading to worse outcome. Solid lines the opposite situation.

A variety of different studies have reported the salutary impact of early T cell reconstitution on post allograft outcomes. The association of T cell recovery kinetics calculated using derivatives of the T cell counts plotted over time, giving the instantaneous rate of change as described here, with transplant relevant clinical outcomes has been reported before^15^. T cell recovery kinetics in that study were associated with GVHD, and survival as in this study. The main mechanism for relapse prevention in RIC allografts is donor immune mediated graft vs. malignancy effect. Historically those who experienced acute & chronic GVHD have had the lowest likelihood of disease relapse^16^, and more recently Grade 2 acute GVHD, but not grade 3-4, has been associated with improved survival and freedom from relapse in both the haploidentical and HLA matched setting^17, 18^. Therefore, in the cohorts presented here, early and intermediate T cell recovery shows strong association with clinical outcomes and may be used to help make decisions about the intensity of immunosuppression employed at different time intervals post-transplant. For example, prolonged immunosuppression in those with very rapid T cell recovery, in the absence of monocyte recovery and *vice versa*, closely following T cell recovery kinetics.

ATG dosing has been evaluated based on AUC achieved following administration^19^. Patients with supra-optimal dosing of ATG in this instance had a higher risk of relapse, while suboptimal exposure led to worsening NRM and worse survival in both groups. In general, lower dose ATG yields superior outcomes following allogeneic transplantation^20^. More intense immunosuppression with alemtuzumab also increases the risk of relapse, with higher mortality seen in these patients^21^. The finding that ATG administered early in the course of conditioning leads to more robust ETIR is consistent with these observations, and indicates that along with dose, optimization of the schedule of administration of ATG is important.

Post-transplant cyclophosphamide (PT-CY) is becoming widely adopted for GVHD prophylaxis following HLA matched^22^ and haploidentical transplantation^23 24^. This modality is associated with significant immune modulation, with different mechanisms of GVHD control proposed^25^. These include an alteration of the Treg-Tcon ratio, delayed T cell recovery kinetics and selective depletion of alloreactive T cell clones^26^. Infections following PT-CY are common, albeit, non-lethal ^27^. ATG when compared with PT-CY retrospectively yields equivalent outcomes in MUD recipients ^28^. Prospective study of ATG vs. PT-CY have failed to identify significant differences in clinical outcomes in single locus HLA mismatched unrelated donor transplants ^29^. Despite the success of PT-CY in expanding the donor pool for patients with hematological malignancy, relapse remains an ongoing problem due to high rates of recurrence of disease in patients thus treated. This may be related to the effective control of alloreactivity, through either T cell clonal deletion or Treg mediated effects. Administration of ATG on a different schedule than the conventional time window of pre-graft infusion is one way to allow adequate immune reconstitution without adversely impacting chronic GVHD risk, and allow dose de-escalation of PT-CY. Indeed, rabbit ATG 2.5 mg/kg given on day -8 thru -5 when added to PT-CY leads to reliable engraftment with no GVHD risk ^30^. Similarly, the ATG schedule reported here leads to improved ETIR without a noticeable impact on overall GVHD risk and with a trend towards improved survival and relapse in the two patient groups studied.

Clinical utility of studying ETIR kinetics aside, this analysis helps understand the physiology at hand. Transplant outcomes are generally modelled as a stochastic process, with probabilities of outcomes varying as a function of certain variables such as conditioning intensity, T cell depletion and GVHD prophylaxis. Modeling immune reconstitution using simple mathematical techniques, and the association of model outputs with clinical outcomes supports the notion that, in aggregate, the immune system behaves as a dynamical system. In dynamical systems, the components of the HCT system, such as, T cells, and antigen presenting cells (monocytes), behave according to predetermined laws. The immune response being proportional to physical properties of the antigen library at hand, such as the number of antigens and their HLA binding affinities, may be one such law. The data here describe the population-wide behavior of classes of immune cells, and likely represents the aggregate T cell clonal behavior, if considered from the molecular, minor histocompatibility antigen-driven perspective. Further in each individual these dynamical T cell responses will be modulated by different cytokine milieu encountered in different individuals. With the expansion in genomic sciences, it is a matter of time before it becomes possible to study these dynamics at the molecular level. The work presented here reinforces the principles identified previously ^31, 32^ that post-transplant immune reconstitution may be mathematically described, and this description may be used to understand the differential impact of different immunosuppressive regimens. This quantitative understanding may also help understand the varying pathophysiology of different GVHD prophylaxis regimens. Thus, while in ATG based regimens, the magnitude of T cell recovery may determine the probability of alloreactivity developing, in PT-CY based regimens this may be a more qualitative relationship, depending on the T cell clonal repertoire specificity, rather than being a simple T cell-magnitude-based association.

Whilst the findings reported here are thought provoking, they need to be confirmed in larger, uniform cohorts of patients. The cohort reported here is relatively small, and varied, as well as the study being retrospective. However, the detailed evaluation of immune reconstitution parameters, from many different, but related angles utilizing a logic-based systematic approach gives confidence in the robustness of the results. In conclusion, quantitative immunobiology of transplantation is reviewed in this paper, by exploring how modification of the schedule of immunosuppressive therapy may modify immune recovery post HCT and effect clinical outcomes. This demonstrates that one may through a deeper understanding of post-transplant immune recovery dynamics be able to ‘fine-tune’ and adjust therapeutic intensity to optimize clinical outcomes.

## Data Availability

Data analyses that supports the findings of this study are available in supplementary figures and tables. Data is not publicly available due to patient privacy restrictions.

## Acknowledgements & author contributions

AT was supported by research funding from the NIH-NCI Cancer Center Support Grant (P30-CA016059; PI: Gordon Ginder, MD). VZ-Designed study, collected and analyzed data, wrote the manuscript; GS-Designed study collected and analyzed data, wrote the manuscript; NR-Collected data, wrote the manuscript; EK-Analyzed data, wrote the manuscript; KH; Critically reviewed and wrote the manuscript; MA-Critically reviewed and wrote the manuscript; JR-Critically reviewed and wrote the manuscript; RG-Critically reviewed and wrote the manuscript; AT-Designed study, directed data analysis, wrote the manuscript.

---

**Supplementary Figure 1.**
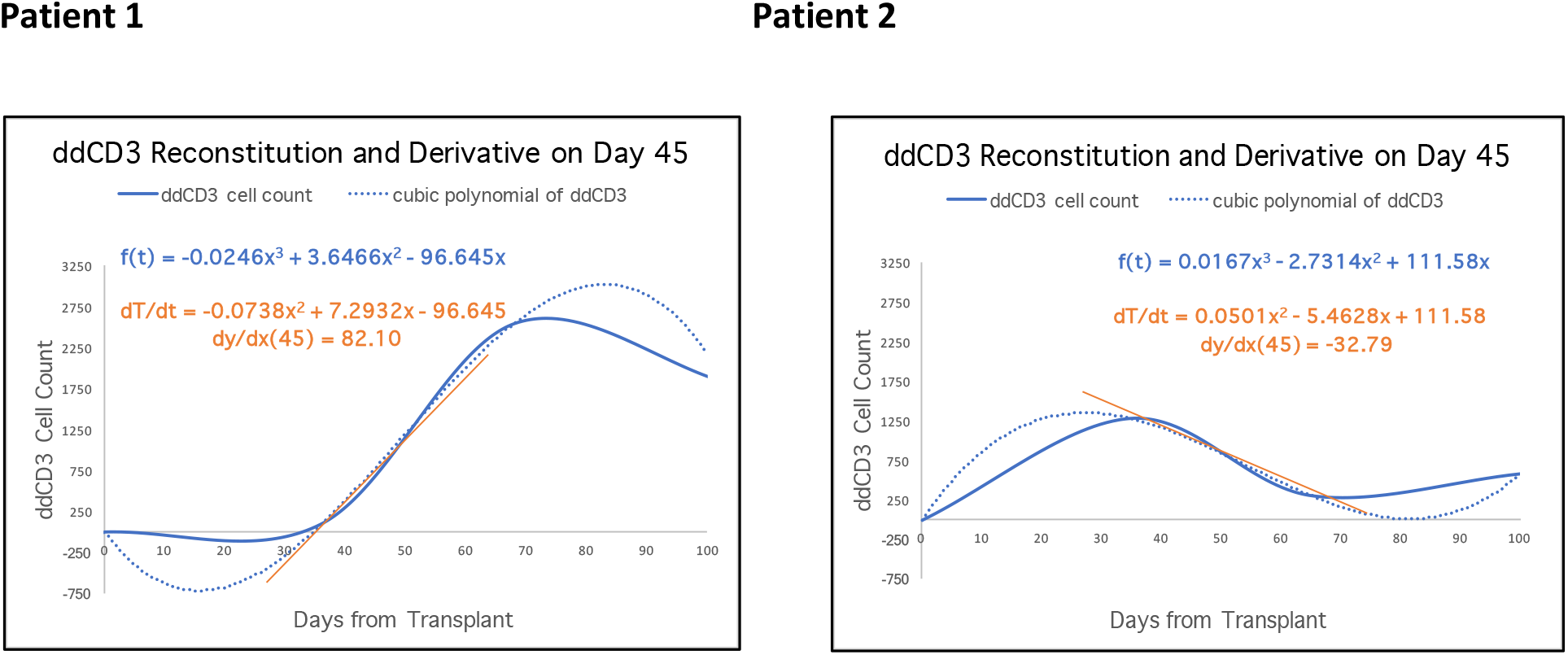
Two patient examples of ddCD3 reconstitution plots. The solid blue line represents the ddCD3 cell count fit to a cubic polynomial and the dotted blue line is its derivative. The solid orange line represents the rate of change at a specific point in time. The two graphs next to each other highlight that while the cell counts on a specific day may be similar, day 45 here, for example, however the rate of recovery may be quite different at that time in the individual patient’s course.

**Supplementary Figure 2.**
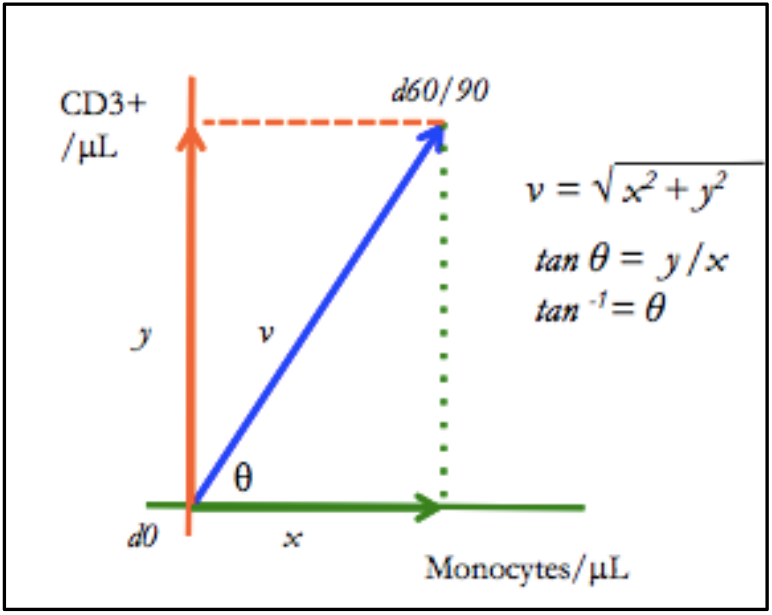
Calculating Monocyte (x-axis) - T cell (y-axis) interaction vector in immune recovery phase space, vector 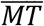. *υ* and *θ* represent vector properties.

**Supplementary Figure 3:**
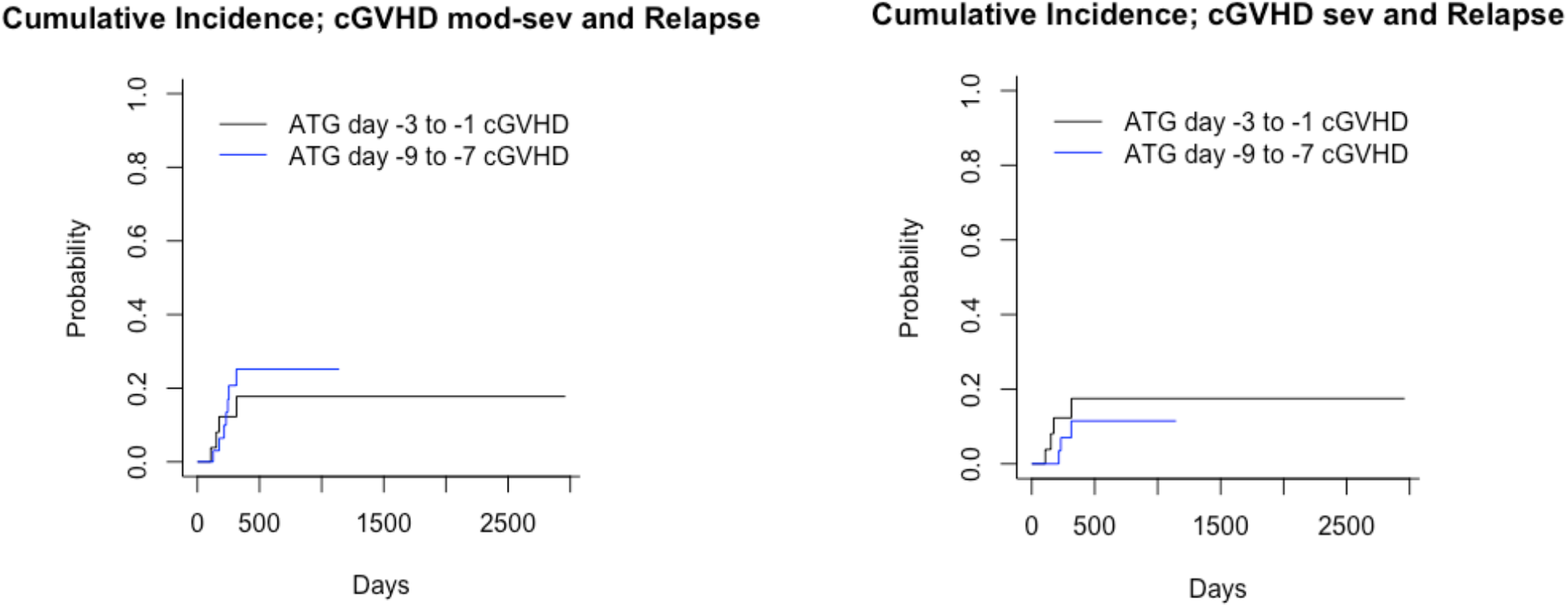
Cumulative Incidence for cGVHD (Grey’s test P=0.63 for moderate to severe and 0.44 for severe). Competing event relapse.

**Supplementary Figure 4.**
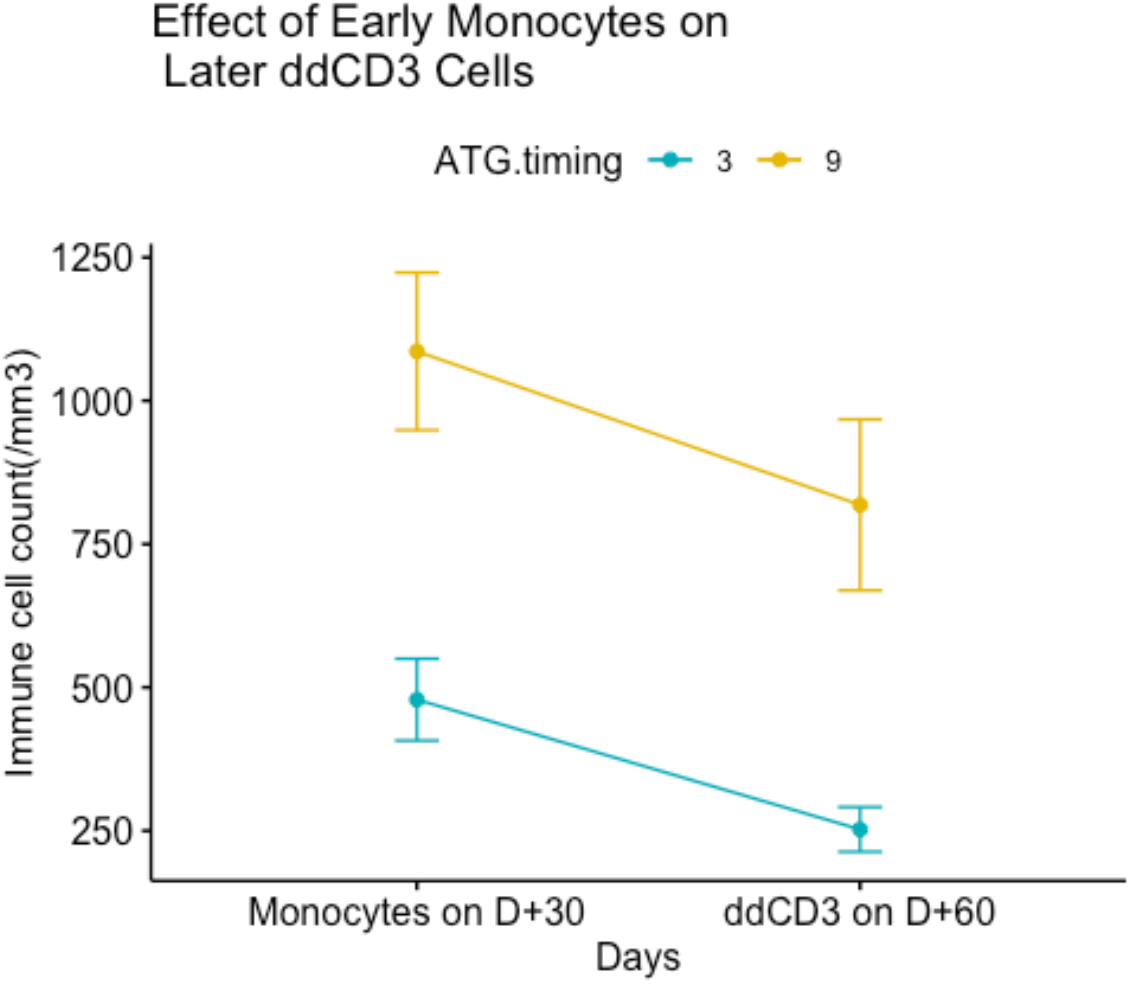
Data separated by ATG cohort, average monocytes on day30 and ddCD3 cells on day 60 (Mean ± S.E.).

**Supplementary Figure 5.**
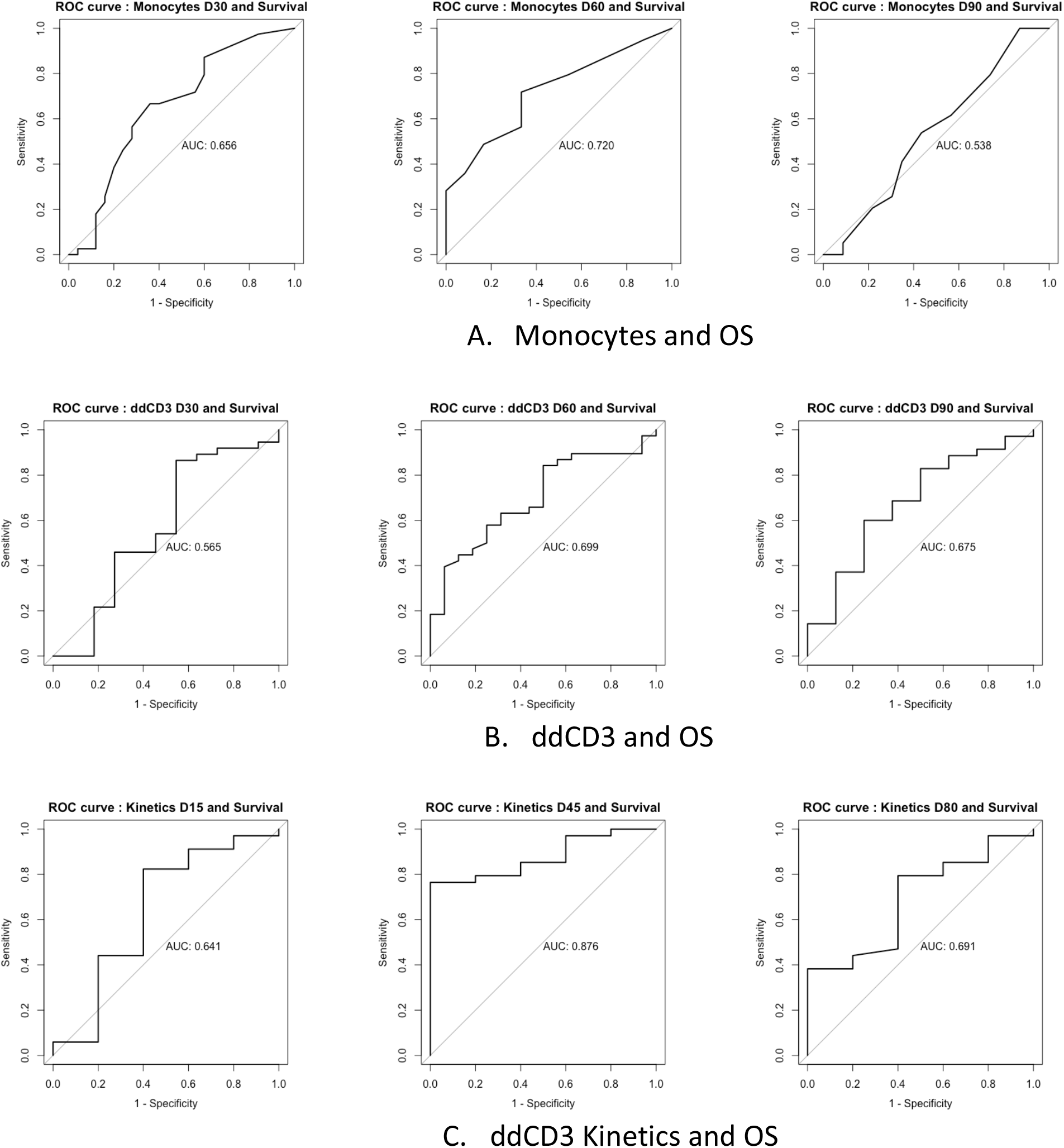
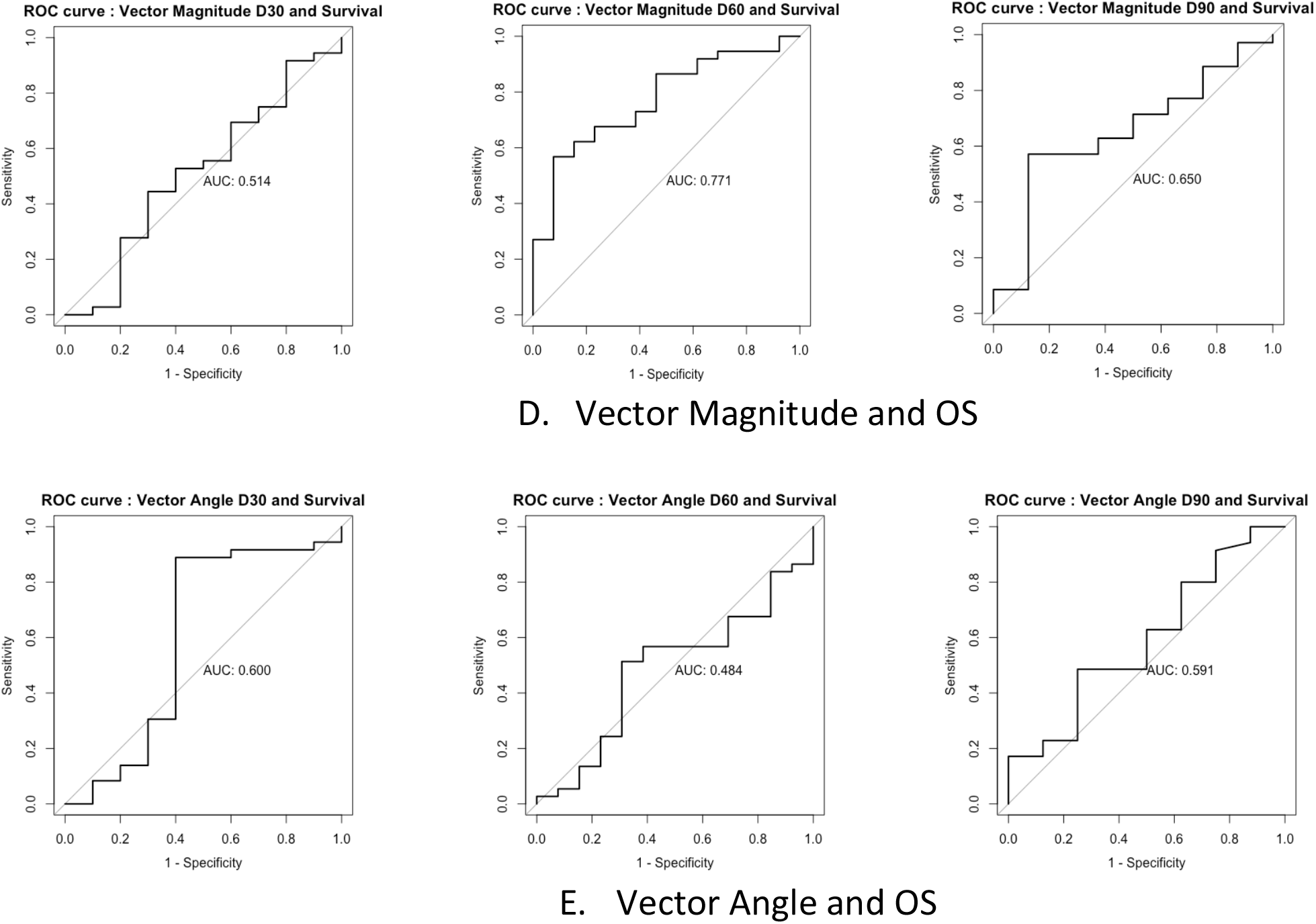
ROC-AUC curves used to determine cutoff for cell counts. Overall survival was used at the outcome in each calculation to determine the value of the cutoff for each day; d30, 60 and 90.

**Supplementary Table 1.** Effect of ATG schedule on clinical outcomes.

| Outcome | Analysis Type | Variable | HR | Lower 95% CI | Upper 95% CI | P |
| --- | --- | --- | --- | --- | --- | --- |
| OS | Multivariate | **ATG timing (-9 vs. -3)** | **0.2508** | **0.0736** | **0.855** | **0.027** |
|  |  | At risk for CMV (Y vs. N) | 0.9672 | 0.2423 | 3.86 | 0.962 |
|  |  | MRD vs. MUD | 1.0905 | 0.287 | 4.144 | 0.899 |
|  |  | Disease Type. Myeloid vs. Lymphoid | 1.8405 | 0.7228 | 4.686 | 0.201 |
|  |  | HLA Match (8/8 vs. 7/8) | 0.3822 | 0.1293 | 1.13 | 0.082 |
|  |  | KPS (100-90 vs. <90) | 0.4788 | 0.1539 | 1.489 | 0.203 |
|  |  | Age at Tx (Recipient) | 1.0556 | 0.9987 | 1.116 | 0.056 |
|  |  | **Sex (F vs. M)** | **3.3613** | **1.1418** | **9.895** | **0.028** |
|  | Univariate | ATG timing (-9 vs. -3) | 0.518 | 0.232 | 1.16 | 0.11 |
| NRM | Multivariate | **ATG timing (-9 vs. -3)** | **0.1442** | **0.0259** | **0.804** | **0.027** |
|  |  | At risk for CMV (Y vs. N) | 0.367 | 0.0596 | 2.259 | 0.28 |
|  |  | MRD vs. MUD | 0.9678 | 0.137 | 6.835 | 0.974 |
|  |  | Disease Type. Myeloid vs. Lymphoid | 0.9002 | 0.203 | 3.992 | 0.89 |
|  |  | **HLA Match (8/8 vs. 7/8)** | **0.1712** | **0.0299** | **0.981** | **0.047** |
|  |  | KPS (100-90 vs. <90) | 0.3703 | 0.0755 | 1.815 | 0.221 |
|  |  | Age at Tx (Recipient) | **1.1457** | **1.0316** | **1.272** | **0.011** |
|  |  | Sex (F vs. M) | 4.2914 | 0.9282 | 19.841 | 0.062 |
|  | Univariate | ATG timing (-9 vs. -3) | 0.505 | 0.16 | 1.59 | 0.24 |
| Relapse | Multivariate | ATG timing (-9 vs. -3) | 0.7773 | 0.181 | 3.35 | 0.735 |
|  |  | At risk for CMV (Y vs. N) | 2.2464 | 0.44 | 11.48 | 0.331 |
|  |  | MRD vs. MUD | 1.3161 | 0.282 | 6.13 | 0.727 |
|  |  | Disease Type. Myeloid vs. Lymphoid | 2.9903 | 0.945 | 9.47 | 0.062 |
|  |  | HLA Match (8/8 vs. 7/8) | 0.5521 | 0.143 | 2.13 | 0.388 |
|  |  | KPS (100-90 vs. <90) | 0.4811 | 0.14 | 1.65 | 0.245 |
|  |  | Age at Tx (Recipient) | 1.0267 | 0.96 | 1.1 | 0.446 |
|  |  | Sex (F vs. M) | 1.6159 | 0.442 | 5.9 | 0.468 |
|  | Univariate | ATG timing (-9 vs. -3) | 0.658 | 0.258 | 1.68 | 0.38 |
| aGVHD 2-4. | Multivariate | ATG timing (-9 vs. -3) | 1.755 | 0.544 | 5.67 | 0.35 |
|  |  | At risk for CMV (Y vs. N) | 0.6511 | 0.208 | 2.04 | 0.46 |
|  |  | MRD vs. MUD | 0.6294 | 0.122 | 3.25 | 0.58 |
|  |  | Disease Type. Myeloid vs. Lymphoid | 0.8815 | 0.279 | 2.79 | 0.83 |
|  |  | HLA Match (8/8 vs. 7/8) | 0.4621 | 0.15 | 1.42 | 0.18 |
|  |  | KPS (100-90 vs. <90) | 1.0661 | 0.394 | 2.88 | 0.9 |
|  |  | Age at Tx (Recipient) | 1.0584 | 0.985 | 1.14 | 0.12 |
|  |  | Sex (F vs. M) | 0.7317 | 0.26 | 2.06 | 0.55 |
|  | Univariate | ATG timing (-9 vs. -3) | 1.579 | 0.669 | 3.73 | 0.3 |
| aGVHD 3-4. | Multivariate | ATG timing (-9 vs. -3) | 0.2754 | 0.0329 | 2.31 | 0.234 |
|  |  | At risk for CMV (Y vs. N) | 0.4898 | 0.0573 | 4.19 | 0.514 |
|  |  | MRD vs. MUD | 0.718 | 0.0631 | 8.16 | 0.789 |
|  |  | Disease Type. Myeloid vs. Lymphoid | 2.3229 | 0.425 | 12.7 | 0.331 |
|  |  | HLA Match (8/8 vs. 7/8) | 0.353 | 0.0559 | 2.23 | 0.268 |
|  |  | KPS (100-90 vs. <90) | 0.3209 | 0.0501 | 2.06 | 0.23 |
|  |  | Age at Tx (Recipient) | 1.0887 | 0.9875 | 1.2 | 0.088 |
|  |  | Sex (F vs. M) | 3.9771 | 0.6438 | 24.57 | 0.137 |
|  | Univariate | ATG timing (-9 vs. -3) | 0.629 | 0.192 | 2.06 | 0.44 |
| cGVHD sev | Multivariate | ATG timing (-9 vs. -3) | 0.5379 | 0.0468 | 6.18 | 0.619 |
|  |  | At risk for CMV (Y vs. N) | 2.3815 | 0.1442 | 39.33 | 0.544 |
|  |  | MRD vs. MUD | 0.426 | 0.0432 | 4.2 | 0.465 |
|  |  | Disease Type. Myeloid vs. Lymphoid | 2.1644 | 0.2785 | 16.82 | 0.46 |
|  |  | HLA Match (8/8 vs. 7/8) | 0.1399 | 0.0172 | 1.14 | 0.066 |
|  |  | KPS (100-90 vs. <90) | 3.7189 | 0.2875 | 48.1 | 0.315 |
|  |  | Age at Tx (Recipient) | 1.0045 | 0.8898 | 1.13 | 0.942 |
|  |  | Sex (F vs. M) | 3.3451 | 0.3614 | 30.96 | 0.288 |
|  | Univariate | ATG timing (-9 vs. -3) | 0.513 | 0.115 | 2.3 | 0.38 |
| cGVHD mod-sev | Multivariate | ATG timing (-9 vs. -3) | 1.46612 | 0.2219 | 9.69 | 0.691 |
|  |  | At risk for CMV (Y vs. N) | 1.3259 | 0.2217 | 7.93 | 0.757 |
|  |  | MRD vs. MUD | 0.49051 | 0.0725 | 3.32 | 0.465 |
|  |  | Disease Type. Myeloid vs. Lymphoid | 1.0089 | 0.1852 | 5.5 | 0.992 |
|  |  | HLA Match (8/8 vs. 7/8) | 0.2428 | 0.0535 | 1.1 | 0.067 |
|  |  | KPS (100-90 vs. <90) | 2.5949 | 0.4736 | 14.22 | 0.272 |
|  |  | Age at Tx (Recipient) | 1.01878 | 0.918 | 1.13 | 0.726 |
|  |  | Sex (F vs. M) | 1.36454 | 0.2849 | 6.54 | 0.697 |
|  | Univariate | ATG timing (-9 vs. -3) | 1.296 | 0.379 | 4.44 | 0.68 |
| RFS | Multivariate | ATG timing (-9 vs. -3) | 0.40818 | 0.133 | 1.25 | 0.117 |
|  |  | At risk for CMV (Y vs. N) | 1.00529 | 0.304 | 3.33 | 0.993 |
|  |  | MRD vs. MUD | 1.12848 | 0.34 | 3.74 | 0.843 |
|  |  | Disease Type. Myeloid vs. Lymphoid | 1.78473 | 0.73 | 4.36 | 0.204 |
|  |  | HLA Match (8/8 vs. 7/8) | 0.37128 | 0.128 | 1.08 | 0.068 |
|  |  | KPS (100-90 vs. <90) | 0.45771 | 0.176 | 1.19 | 0.109 |
|  |  | Age at Tx (Recipient) | 1.06889 | 1.007 | 1.13 | 0.029 |
|  |  | Sex (F vs. M) | 2.57274 | 0.967 | 6.84 | 0.058 |
|  | Univariate | ATG timing (-9 vs. -3) | 0.591 | 0.268 | 1.22 | 0.16 |

**Supplementary Table 2.**
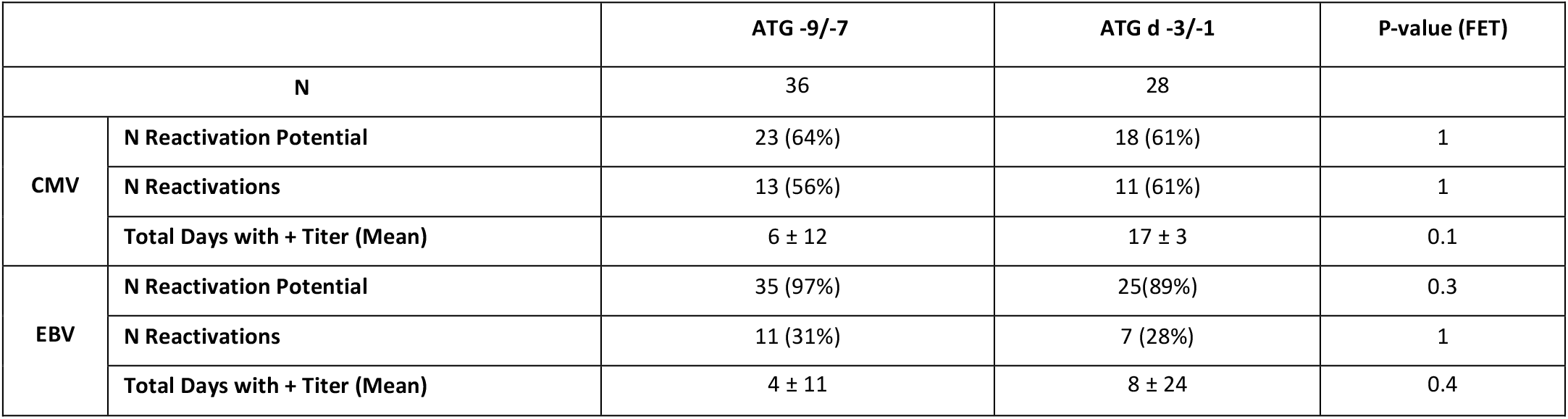
EBV and CMV reactivation in the two cohorts. Patients receiving Letermovir (n=4) removed from this analysis. In the analysis of ‘Total Days with + Titers’, a cut off of 180 days of follow-up was used, and patients without clinically significant reactivation (CMV >500/EBV >100 copies) included.

**Supplementary Table 3.**
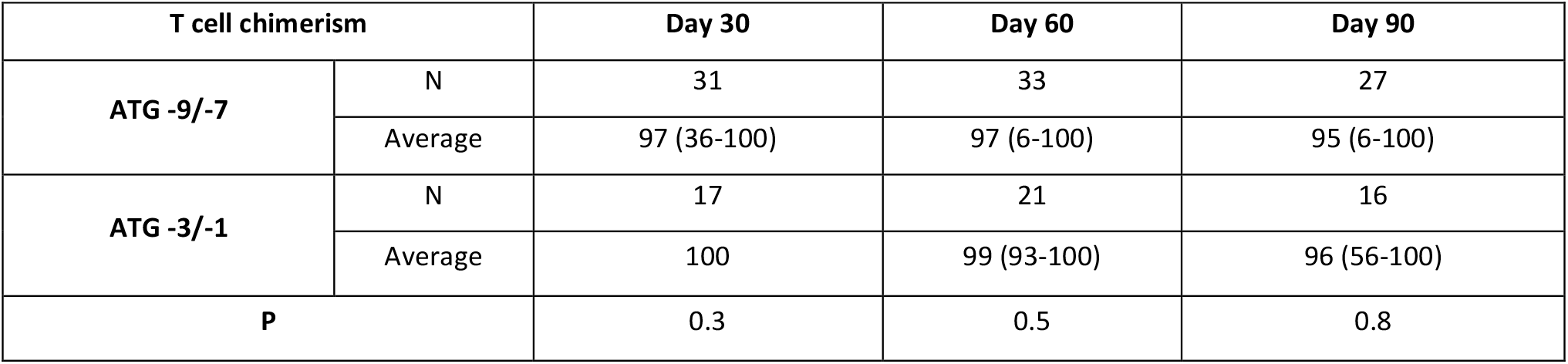
T cell Chimerism following SCT at days 30, 60, 90.

| T cell chimerism |  | Day 30 | Day 60 | Day 90 |
| --- | --- | --- | --- | --- |
| ATG -9/-7 | N | 31 | 33 | 27 |
|  | Average | 97 (36-100) | 97 (6-100) | 95 (6-100) |
| ATG -3/-1 | N | 17 | 21 | 16 |
|  | Average | 100 | 99 (93-100) | 96 (56-100) |
| P |  | 0.3 | 0.5 | 0.8 |

**Supplementary Table 4.**
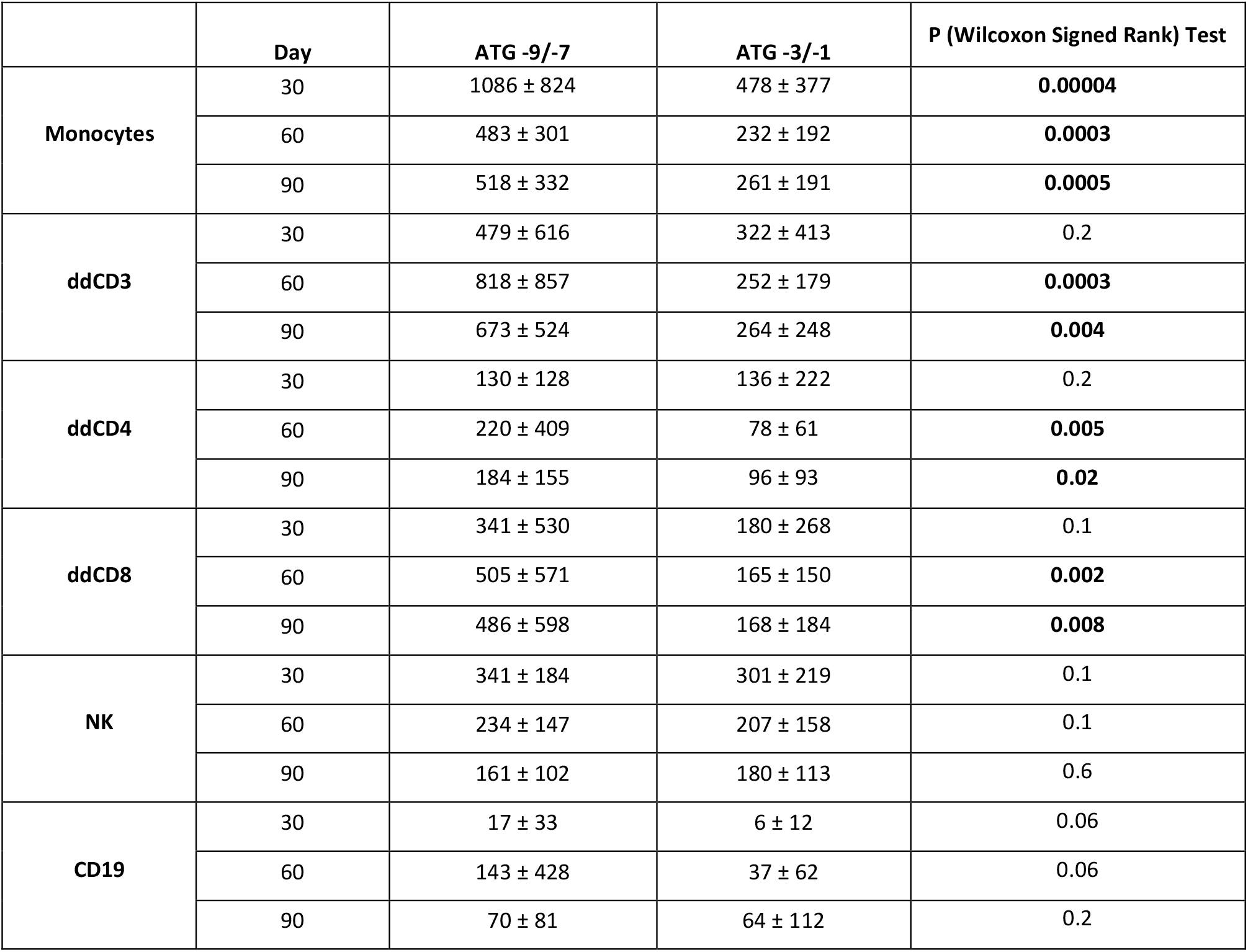
Absolute cell count recovery following allogeneic HCT in the two cohorts

**Supplementary Table 5A.**
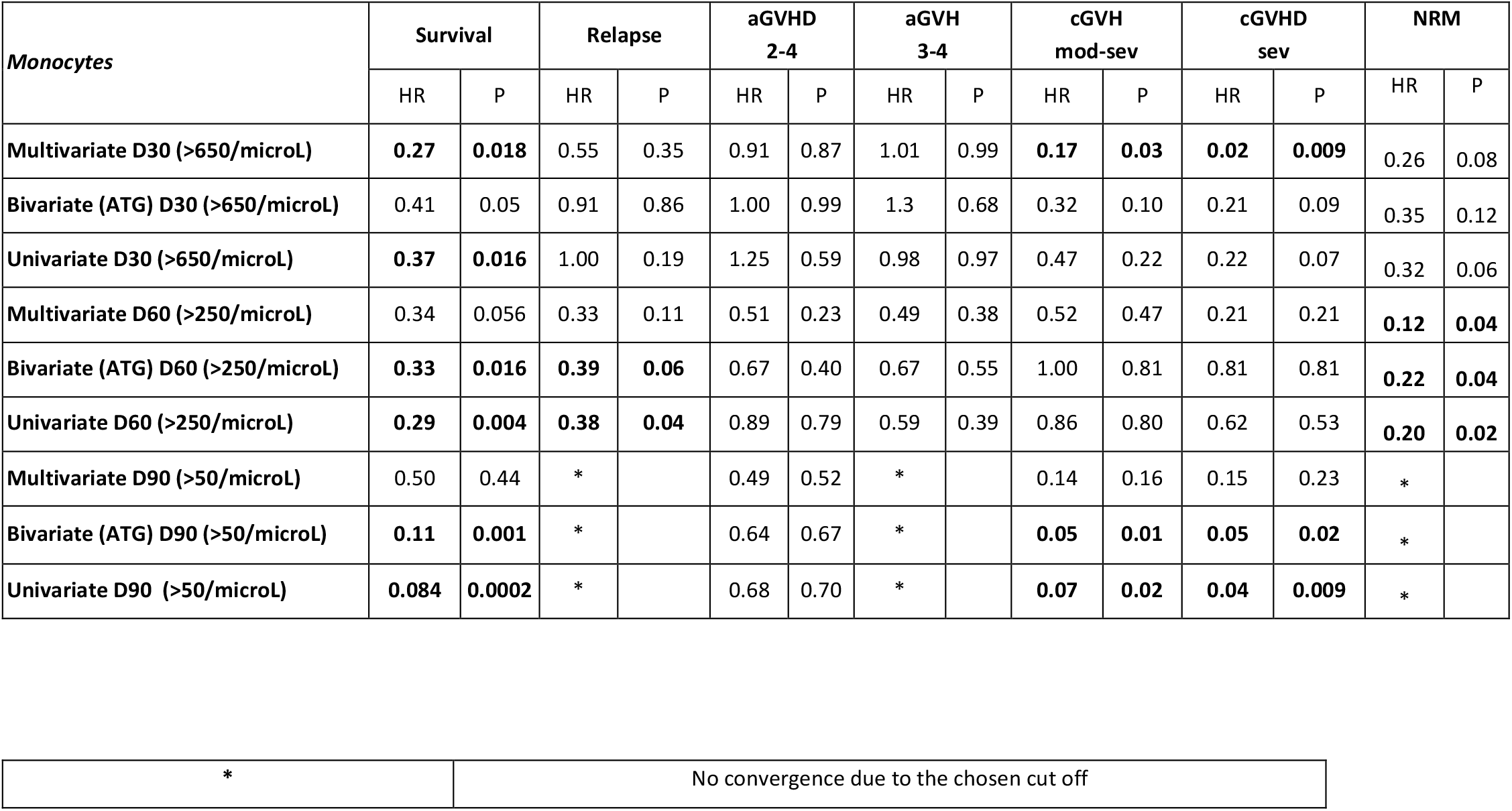
Impact of monocyte count recovery on clinical outcomes

| Monocytes | Survival |  | Relapse |  | aGVHD 2-4 |  | aGVH 3-4 |  | cGVH mod-sev |  | cGVHD sev |  | NRM |  |
| --- | --- | --- | --- | --- | --- | --- | --- | --- | --- | --- | --- | --- | --- | --- |
|  | HR | P | HR | P | HR | P | HR | P | HR | P | HR | P | HR | P |
| Multivariate D30 (>650/microL) | **0.27** | **0.018** | 0.55 | 0.35 | 0.91 | 0.87 | 1.01 | 0.99 | **0.17** | **0.03** | **0.02** | **0.009** | 0.26 | 0.08 |
| Bivariate (ATG) D30 (>650/microL) | 0.41 | 0.05 | 0.91 | 0.86 | 1.00 | 0.99 | 1.3 | 0.68 | 0.32 | 0.10 | 0.21 | 0.09 | 0.35 | 0.12 |
| Univariate D30 (>650/microL) | **0.37** | **0.016** | 1.00 | 0.19 | 1.25 | 0.59 | 0.98 | 0.97 | 0.47 | 0.22 | 0.22 | 0.07 | 0.32 | 0.06 |
| Multivariate D60 (>250/microL) | 0.34 | 0.056 | 0.33 | 0.11 | 0.51 | 0.23 | 0.49 | 0.38 | 0.52 | 0.47 | 0.21 | 0.21 | **0.12** | **0.04** |
| Bivariate (ATG) D60 (>250/microL) | **0.33** | **0.016** | **0.39** | **0.06** | 0.67 | 0.40 | 0.67 | 0.55 | 1.00 | 0.81 | 0.81 | 0.81 | **0.22** | **0.04** |
| Univariate D60 (>250/microL) | **0.29** | **0.004** | **0.38** | **0.04** | 0.89 | 0.79 | 0.59 | 0.39 | 0.86 | 0.80 | 0.62 | 0.53 | **0.20** | **0.02** |
| Multivariate D90 (>50/microL) | 0.50 | 0.44 | * |  | 0.49 | 0.52 | * |  | 0.14 | 0.16 | 0.15 | 0.23 | * |  |
| Bivariate (ATG) D90 (>50/microL) | **0.11** | **0.001** | * |  | 0.64 | 0.67 | * |  | **0.05** | **0.01** | **0.05** | **0.02** | * |  |
| Univariate D90 (>50/microL) | **0.084** | **0.0002** | * |  | 0.68 | 0.70 | * |  | **0.07** | **0.02** | **0.04** | **0.009** | * |  |

|  |  |
| --- | --- |
| * | No convergence due to the chosen cut off |

**Supplementary Table 5B.**
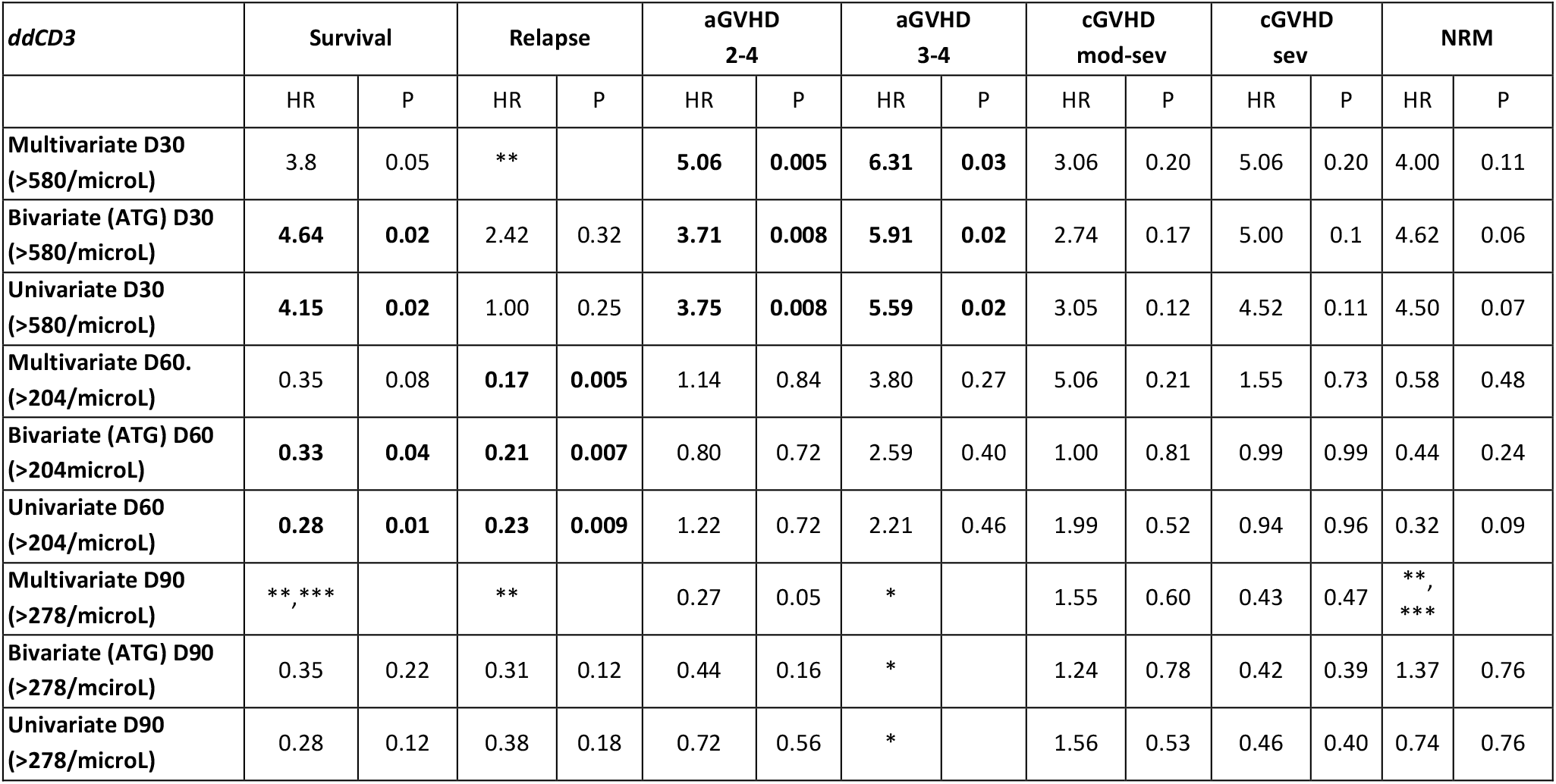
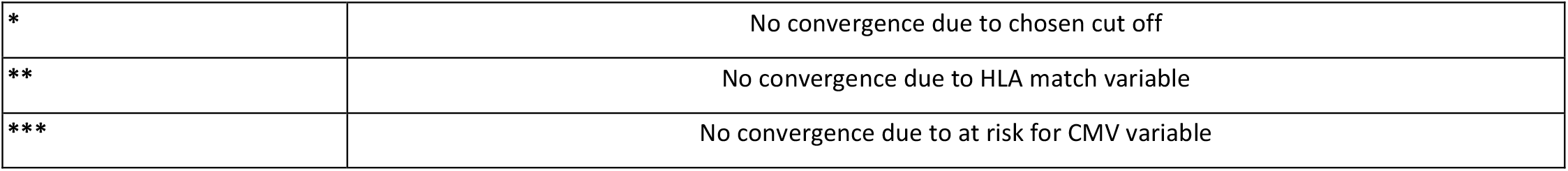
Impact of donor derived T cell recovery on clinical outcomes.

| ddCD3 | Survival |  | Relapse |  | aGVHD 2-4 |  | aGVHD 3-4 |  | cGVHD mod-sev |  | cGVHD sev |  | NRM |  |
| --- | --- | --- | --- | --- | --- | --- | --- | --- | --- | --- | --- | --- | --- | --- |
|  | HR | P | HR | P | HR | P | HR | P | HR | P | HR | P | HR | P |
| Multivariate D30 (>580/microL) | 3.8 | 0.05 | ** |  | **5.06** | **0.005** | **6.31** | **0.03** | 3.06 | 0.20 | 5.06 | 0.20 | 4.00 | 0.11 |
| Bivariate (ATG) D30 (>580/microL) | **4.64** | **0.02** | 2.42 | 0.32 | **3.71** | **0.008** | **5.91** | **0.02** | 2.74 | 0.17 | 5.00 | 0.1 | 4.62 | 0.06 |
| Univariate D30 (>580/microL) | **4.15** | **0.02** | 1.00 | 0.25 | **3.75** | **0.008** | **5.59** | **0.02** | 3.05 | 0.12 | 4.52 | 0.11 | 4.50 | 0.07 |
| Multivariate D60. (>204/microL) | 0.35 | 0.08 | **0.17** | **0.005** | 1.14 | 0.84 | 3.80 | 0.27 | 5.06 | 0.21 | 1.55 | 0.73 | 0.58 | 0.48 |
| Bivariate (ATG) D60 (>204microL) | **0.33** | **0.04** | **0.21** | **0.007** | 0.80 | 0.72 | 2.59 | 0.40 | 1.00 | 0.81 | 0.99 | 0.99 | 0.44 | 0.24 |
| Univariate D60 (>204/microL) | **0.28** | **0.01** | **0.23** | **0.009** | 1.22 | 0.72 | 2.21 | 0.46 | 1.99 | 0.52 | 0.94 | 0.96 | 0.32 | 0.09 |
| Multivariate D90 (>278/microL) | ** , *** |  | ** |  | 0.27 | 0.05 | * |  | 1.55 | 0.60 | 0.43 | 0.47 | **, *** |  |
| Bivariate (ATG) D90 (>278/microL) | 0.35 | 0.22 | 0.31 | 0.12 | 0.44 | 0.16 | * |  | 1.24 | 0.78 | 0.42 | 0.39 | 1.37 | 0.76 |
| Univariate D90 (>278/microL) | 0.28 | 0.12 | 0.38 | 0.18 | 0.72 | 0.56 | * |  | 1.56 | 0.53 | 0.46 | 0.40 | 0.74 | 0.76 |

|  |  |
| --- | --- |
| * | No convergence due to chosen cut off |
| ** | No convergence due to HLA match variable |
| *** | No convergence due to at risk for CMV variable |

**Supplementary Table 6.** Rate of change of T cell recovery between the two cohorts (cells/μL • day^−1^).

| *Rate of change (day)* | **ATG -9/-7 (N=26)** | **ATG -3/-1 (N=13)** | **P (Wilcoxon)** |
| --- | --- | --- | --- |
| **dT/dt (15)** | 16.7 ± 21.9 | 8.2 ± 10.8 | **0.03** |
| **dT/dt (45)** | 14.2 ± 38.8 | 0.5 ± 10.6 | 0.3 |
| **dT/dt (80)** | -4.3 ± 10.5 | 0.2 ± 3.6 | 1 |

**Supplementary Table 7.**
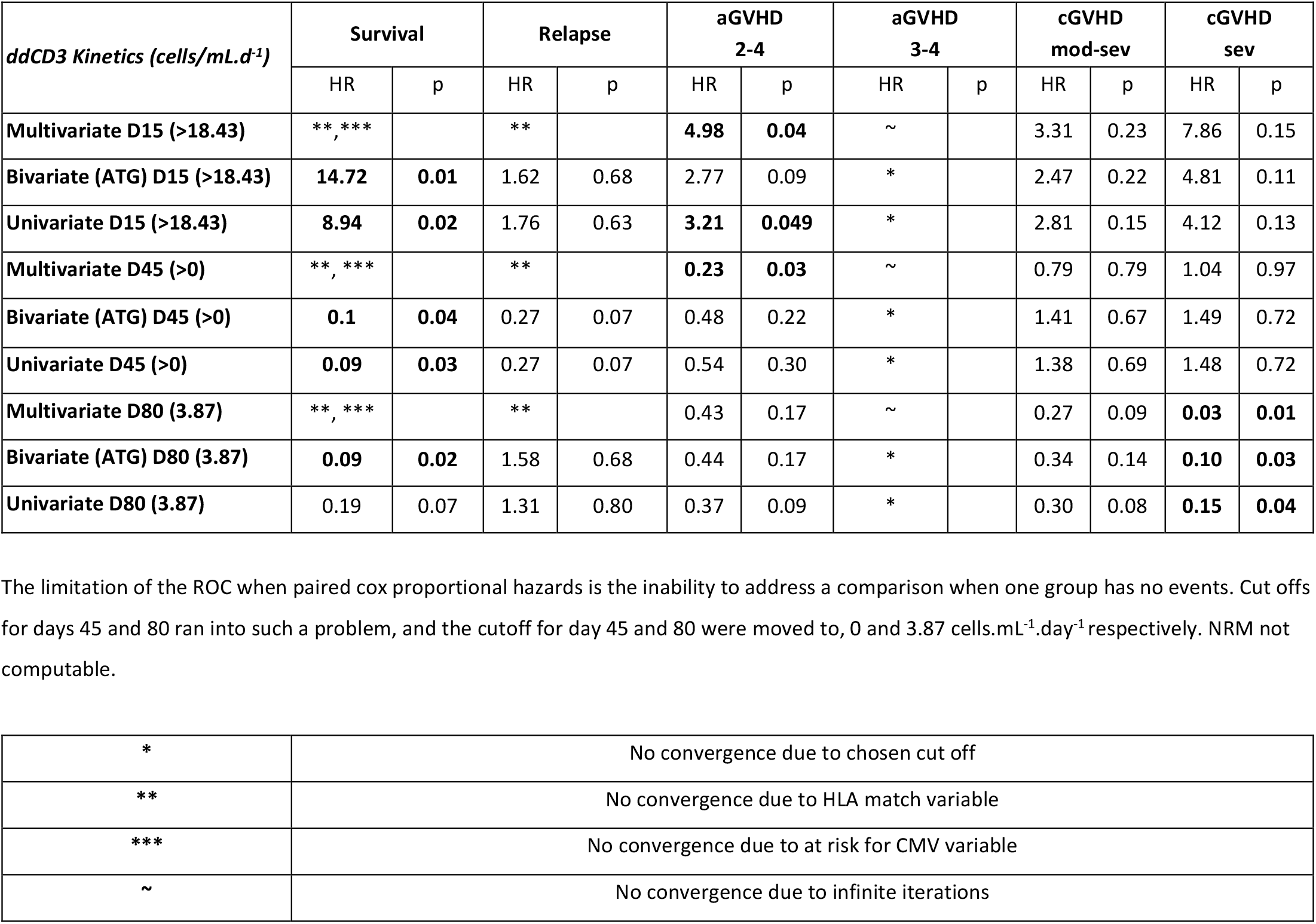
The T cell kinetics analyses, looking at OS, relapse, and GVHD.

| *ddCD3 Kinetics (cells/mL.d<sup>-1</sup>)* | Survival |  | Relapse |  | aGVHD 2-4 |  | aGVHD 3-4 |  | cGVHD mod-sev |  | cGVHD sev |  |
| --- | --- | --- | --- | --- | --- | --- | --- | --- | --- | --- | --- | --- |
|  | HR | p | HR | p | HR | p | HR | p | HR | p | HR | p |
| **Multivariate D15 (>18.43)** | **\*\* , \*\*\*** |  | **\*\*** |  | **4.98** | **0.04** | ~ |  | 3.31 | 0.23 | 7.86 | 0.15 |
| **Bivariate (ATG) D15 (>18.43)** | **14.72** | **0.01** | 1.62 | 0.68 | 2.77 | 0.09 | * |  | 2.47 | 0.22 | 4.81 | 0.11 |
| **Univariate D15 (>18.43)** | **8.94** | **0.02** | 1.76 | 0.63 | **3.21** | **0.049** | * |  | 2.81 | 0.15 | 4.12 | 0.13 |
| **Multivariate D45 (>0)** | **\*\* , \*\*\*** |  | **\*\*** |  | **0.23** | **0.03** | ~ |  | 0.79 | 0.79 | 1.04 | 0.97 |
| **Bivariate (ATG) D45 (>0)** | **0.1** | **0.04** | 0.27 | 0.07 | 0.48 | 0.22 | * |  | 1.41 | 0.67 | 1.49 | 0.72 |
| **Univariate D45 (>0)** | **0.09** | **0.03** | 0.27 | 0.07 | 0.54 | 0.30 | * |  | 1.38 | 0.69 | 1.48 | 0.72 |
| **Multivariate D80 (3.87)** | **\*\* , \*\*\*** |  | **\*\*** |  | 0.43 | 0.17 | ~ |  | 0.27 | 0.09 | **0.03** | **0.01** |
| **Bivariate (ATG) D80 (3.87)** | **0.09** | **0.02** | 1.58 | 0.68 | 0.44 | 0.17 | * |  | 0.34 | 0.14 | **0.10** | **0.03** |
| **Univariate D80 (3.87)** | 0.19 | 0.07 | 1.31 | 0.80 | 0.37 | 0.09 | * |  | 0.30 | 0.08 | **0.15** | **0.04** |

|  |  |
| --- | --- |
| * | No convergence due to chosen cut off |
| ** | No convergence due to HLA match variable |
| *** | No convergence due to at risk for CMV variable |
| ~ | No convergence due to infinite iterations |

**Supplementary Table 8.** Monocyte-T cell immune recovery vector (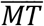) in the two ATG cohorts.

| *Vector Component* | Day | ATG -9/-7 | ATG -3/-1 | p-value (Wilcoxon) |
| --- | --- | --- | --- | --- |
| Magnitude | 30 | 1218 ± 601 | 717 ± 519 | **0.005** |
|  | 60 | 1087 ± 809 | 343 ± 184 | **<0.0001** |
|  | 90 | 887 ± 476 | 365 ± 238 | **<0.0001** |
| Angle | 30 | 23 ± 24 | 27 ± 25 | 0.7 |
|  | 60 | 48 ± 27 | 48 ± 23 | 0.5 |
|  | 90 | 48 ± 24 | 44 ± 25 | 0.3 |

**Supplementary Table 9.** Immune vectors properties and clinical outcomes. The vector (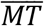) analyses, looking at OS, relapse, and GVHD.

| Vector Magnitude | Survival |  | Relapse |  | aGVHD 2-4 |  | aGVHD 3-4 |  | cGVHD mod-sev |  | cGVHD sev |  | NRM |  |
| --- | --- | --- | --- | --- | --- | --- | --- | --- | --- | --- | --- | --- | --- | --- |
|  | HR | p | HR | p | HR | p | HR | p | HR | p | HR | p | HR | p |
| Multivariate D30 (>955.25) | 2.53 | 0.25 | ** |  | 1.33 | 0.64 | 3.52 | 0.23 | 2.12 | 0.41 | 0.32 | 0.31 | 2.87 | 0.38 |
| Bivariate (ATG) D30 (>955.25) | 3.38 | 0.11 | 7.90 | 0.06 | 1.30 | 0.65 | 3.65 | 0.15 | 1.71 | 0.51 | 0.61 | 0.64 | 3.20 | 0.24 |
| Univariate D30 (>955.25) | 1.91 | 0.35 | 1.76 | 0.63 | 1.35 | 0.56 | 1.75 | 0.50 | 2.0 | 0.33 | 0.60 | 0.58 | 1.61 | 0.58 |
| Multivariate D60 (>619) | **0.08** | **0.03** | 0.43 | 0.35 | 0.83 | 0.78 | *, *** |  | 1.48 | 0.70 | 0.26 | 0.40 | 0.14 | 0.12 |
| Bivariate (ATG) D60 (>619) | **0.09** | **0.03** | 0.47 | 0.37 | 0.9 | 0.86 | * |  | 1.00 | 0.81 | 0.63 | 0.69 | 0.17 | 0.14 |
| Univariate D60 (>619) | **0.09** | **0.02** | 0.59 | 0.45 | 1.346 | 0.55 | * |  | 2.15 | 0.28 | 0.67 | 0.66 | 0.13 | 0.05 |
| Multivariate D90 (>643.05) | **, *** |  | ** |  | 0.94 | 0.93 | * |  | 0.87 | 0.87 | 0.04 | 0.07 | 1.02 | 0.99 |
| Bivariate (ATG) D90 (>643.05) | 0.17 | 0.11 | 0.41 | 0.24 | 1.18 | 0.79 | * |  | 0.86 | 0.84 | 0.16 | 0.12 | 0.57 | 0.66 |
| Univariate D90 (>643.05) | 0.14 | 0.07 | 0.50 | 0.32 | 1.87 | 0.27 | * |  | 1.18 | 0.80 | 0.21 | 0.17 | 0.30 | 0.30 |
| Vector Angle |  |  |  |  |  |  |  |  |  |  |  |  |  |  |
| Multivariate D30 (>34.65) | 9.96 | 0.007 | ** |  | 4.52 | 0.006 | 16.38 | 0.01 | 9.15 | 0.006 | *, **, *** |  | 12.05 | 0.019 |
| Bivariate (ATG) D30 (>34.65) | 8.72 | 0.001 | 3.14 | 0.20 | 4.00 | 0.005 | 11.02 | 0.004 | 6.79 | 0.007 | 18.34 | 0.004 | 9.7 | 0.009 |
| Univariate D30 (>34.65) | 8.15 | 0.001 | 1.76 | 0.63 | 4.00 | 0.006 | 11.37 | 0.004 | 7.24 | 0.006 | 15.00 | 0.005 | 10.02 | 0.008 |
| Multivariate D60 (>49.7) | 2.28 | 0.21 | 0.15 | 0.03 | 0.90 | 0.83 | 3.35 | 0.28 | 2.15 | 0.31 | 1.73 | 0.57 | 3.60 | 0.13 |
| Bivariate (ATG) D60 (>49.7) | 1.95 | 0.27 | 0.40 | 0.19 | 0.85 | 0.75 | 3.25 | 0.29 | 1.00 | 0.81 | 1.31 | 0.77 | 2.87 | 0.19 |
| Univariate D60 (>49.7) | 2.05 | 0.23 | 0.40 | 0.19 | 0.82 | 0.70 | 3.56 | 0.26 | 1.83 | 0.39 | 1.29 | 0.78 | 3.10 | 0.16 |
| Multivariate D90 (>55) | **, *** |  | ~ |  | 0.5774 | 0.35 | **, *** |  | 1.53 | 0.59 | 4.07 | 0.24 | 0.65 | 0.71 |
| Bivariate (ATG) D90 (>55) | 0.47 | 0.36 | * |  | 0.68 | 0.50 | 1.40 | 0.81 | 1.25 | 0.74 | 1.75 | 0.55 | 1.40 | 0.74 |
| Univariate D90 (>55) | 0.42 | 0.29 | * |  | 0.80 | 0.69 | 1.30 | 0.85 | 1.39 | 0.62 | 1.68 | 0.57 | 1.16 | 0.88 |

